# Differentiating benign from malignant adnexal masses by biomarker-agnostic plasma proteomics using adaptive machine learning

**DOI:** 10.64898/2026.09.23.26363813

**Authors:** Johannes B. Müller-Reif, Sarah Ackroyd, Vincent Albrecht, Melanie Weigert, Kathrin Korff, Alicia-Sophie Schebesta, Roni Yoeli-Bik, Maryam Siddiqui, Andrew Rausch, Emese Zsiros, Matthias Mann, Ernst Lengyel

**Author notes:** equal contribution (Johannes B. Müller-Reif, Sarah Ackroyd, Matthias Mann, Ernst Lengyel).

## Abstract

**Background:** Pre-operative triage of adnexal masses with serum CA-125, HE4, and ultrasound (O-RADS) has limited accuracy, contributing to unnecessary surgery. We develop and validate a first of its kind, biomarker panel-free plasma proteomic classifier that distinguishes malignant from benign adnexal masses from a single blood draw, operating directly on discovery-mode mass spectrometry.

**Methods:** In this multicenter prospective observational study, plasma from women with adnexal masses and from healthy controls at a large urban academic center (discovery and four validation cohorts) and an external academic center from another Urban-suburban area (external validation) was analyzed by data-independent-acquisition mass spectrometry (Orbitrap Astral), quantifying >1,000 proteins per sample. Cancerous (malignant, borderline adnexal mass, or metastasis to ovary) versus benign adnexal masses were classified with ADAPT-MS (Adaptive Diagnostic Architecture for Personalized Testing by Mass Spectrometry), which retrains an ElasticNet model for each sample based on the proteins measured. The primary outcome was discrimination (area under the receiver operating characteristic curve [AUC]). Pre-operative O-RADS scores and serum CA-125 were compared and combined with proteomics on matched subsets.

**Findings:** Using a discovery cohort (n=1062), an ADAPT-MS classifier, validated across five independent cohorts (n=623), achieved a summary AUC of 0.835 (cohort range 0.779–0.904). Among patients with pre-operative O-RADS (n=231), proteomics (AUC 0.852) was comparable to O-RADS alone (0.863) and combining them raised discrimination to 0.922. Proteomics outperformed serum CA-125 (0.915 versus 0.777; n=125) and proteome-derived ROMA-like and two-marker surrogates. Biologically, the classification-relevant proteins are enriched for host response proteins rather than tumor cell associated protein changes, comprising ECM remodeling, complement and innate immune response pathways. This explains good detection of tumor with heterogenous pathology and also metastasis to ovary.

**Interpretation:** A single plasma proteomic measurement interpreted using adaptive machine learning discriminates malignant from benign adnexal masses with good internal and external validation, and is complementary to ultrasound, warranting prospective evaluation as a panel-free diagnostic adjunct to imaging.

**Funding:** Funded by the Max Planck Society for the Advancement of Sciences (MM), an NIH R35 grant (EL), and support from Arthur and Nicole Herbst (EL).

**Research in context:** *Evidence before this study:* Presence of protein biomarkers for ovarian cancer and in detail, adnexal mass triage is historically present e.g. by CA-125 or HE4. Proteomics studies have shown many more potential protein markers that might be used as biomarkers instead or additional to the presently used ones. Classification algorithms however do currently rely on single or few biomarkers only, limiting sensitivity and specificity.

*Added value of this study:* To our knowledge, this study is the first example of a proteomics classifier that uses ML on discovery-based MS-proteomics data at the single-sample level and thus has the potential for diagnostic use. While in the past, proteomics has been used as a discovery tool and potential markers needed to be subjected to a follow-up assay development, we show by application to 5 different independent validation cohorts that a proteomics classifier based on the recently developed architecture Adaptive Diagnostic Architecture for Personalized Testing by Mass Spectrometry (ADAPT-MS) generalizes in performance between discovery and validation cohorts and can be applied at the single sample level.

*Implications of all the available evidence:* We demonstrate that proteomics can be applied beyond discovery and classify adnexal masses together with ultrasound at an AUC of 0.922. This sets a precedence case for other diagnostic applications of proteomics, both in study design (plasma proteomics is always measured and used to best of ability in discovery and validation), and in single-sample application mode by ADAPT-MS.

## INTRODUCTION

Ovarian cancer is the leading cause of death from gynecological malignancy, with an estimated 21,000 (44,000) new cases and 14,500 (30,000) deaths in the USA (EU) (Siegel et al. 2026; Ferlay et al. 2019). Prognosis for high-grade serous ovarian cancer is dependent on stage at diagnosis, with five-year overall survival exceeding 90% for disease detected at stage I but falling below 50-60% for stage III–IV disease (Kurnit, Fleming, and Lengyel 2021). Because most women present with advanced disease, the early and accurate identification of malignant adnexal masses is a significant unmet need.

Adnexal masses are common, occurring in approximately one in five women during their lifetime, with the great majority being benign—functional cysts, endometriomas, teratomas, and cystadenomas. The clinical task is therefore to distinguish the minority of malignant masses that warrant prompt referral to gynecological oncology from benign lesions suitable for conservative (non-surgical) or general gynecological management (‘Practice Bulletin No. 174: Evaluation and Management of Adnexal Masses’ 2016) Reaffirmed in 2023. Current triage integrates clinical assessment, imaging, and serum biomarkers. Transvaginal ultrasound is the primary imaging modality, and structured systems—the ADNEX (Assessment of Different NEoplasias in the adneXa) model and the American College of Radiology Ovarian-Adnexal Reporting and Data System (O-RADS)—provide reproducible risk estimates based on expertly measured ultrasound features (Andreotti et al. 2020; Yoeli-Bik et al. 2023). However, these systems have reduced accuracy in early-stage, non-epithelial disease, and depend on specialist sonographic expertise, expert radiologist review, and expensive ultrasound equipment that may not be readily available in all settings and is highly operator-dependent.

Serum CA-125 (*MUC16 gene*) is currently the best single-protein biomarker to differentiate benign from malignant adnexal masses, but it has limited sensitivity, particularly for early-stage malignancy and non-serous histologies (rarer but representing 10% of cases), and limited specificity because it is elevated in many benign conditions, including inflammation-mediated conditions such as endometriosis, peritonitis, and cirrhosis (Bast et al. 1983; Babic et al. 2017). Human epididymis protein 4 (HE4, the product of the *WFDC2* gene) and the Risk of Ovarian Malignancy Algorithm (ROMA), which combines CA-125 and HE4 with menopausal status, improve prediction of malignancy but, because of their low specificity, remain insufficient for definitive triage of an adnexal mass (Davenport et al. 2022). The OVA1^®^ and OVERO^®^ multivariate index assays, which measure five serum proteins, achieved FDA clearance but have low specificity (36-71%) and sensitivity (89-92%) (Ueland et al. 2011; Fritsche and Bullock 2023). None of these methods provide a comprehensive profile of the circulating proteome; therefore, we hypothesized that a broad proteomic analysis could help distinguish benign from malignant adnexal masses.

Mass spectrometry (MS)-based plasma proteomics offers an alternative to single-protein biomarkers by quantifying thousands of proteins in a single measurement and capturing the systemic molecular response to malignancy (Aebersold and Mann 2016; Bader, Albrecht, and Mann 2023). Advances in sample preparation and instrumentation, including the PCA-N workflow and the Orbitrap Astral analyzer, have made deep plasma profiling feasible at clinical-scale throughput (Albrecht et al. 2025; Guo, Steen, and Mann 2025). Yet translation has been limited by missing values in individual samples, the need to develop targeted assays, and the difficulty of applying a single fixed panel across heterogeneous patients (Rifai, Gillette, and Carr 2006). We previously developed a machine learning application, Adaptive Diagnostic Architecture for Personalized Testing by Mass Spectrometry (ADAPT-MS), which overcomes these limitations by retraining classifiers on the proteins actually measured in each sample, and validated it across sepsis, metabolic syndrome, and Alzheimer’s disease (Müller-Reif et al. 2026).

By applying ADAPT-MS to adnexal-mass classification, we showcase a novel approach utilizing discovery proteomics and panel-free adaptive classification specifically for adnexal mass triage. Using a discovery cohort and five validation cohorts— including one from a different institution—we demonstrate that plasma proteomics can effectively serve as a practical diagnostic tool to distinguish adnexal masses. This method enhances and complements existing diagnostic tools like ultrasound (O-RADS) and serum biomarkers (CA-125, HE4) commonly used in routine care, achieving high accuracy in identifying malignancies.

## RESULTS

### Cohort characteristics

The study design, which addresses whether a proteomic signature can differentiate benign from malignant adnexal masses, is shown in Figure 1. Conceptually, we used a large discovery cohort, confirmed results with 4 internal and 1 external validation cohorts, employed the latest liquid chromatography-mass spectrometry methodology, and followed with adaptive machine-learning analysis.

**Figure 1.**
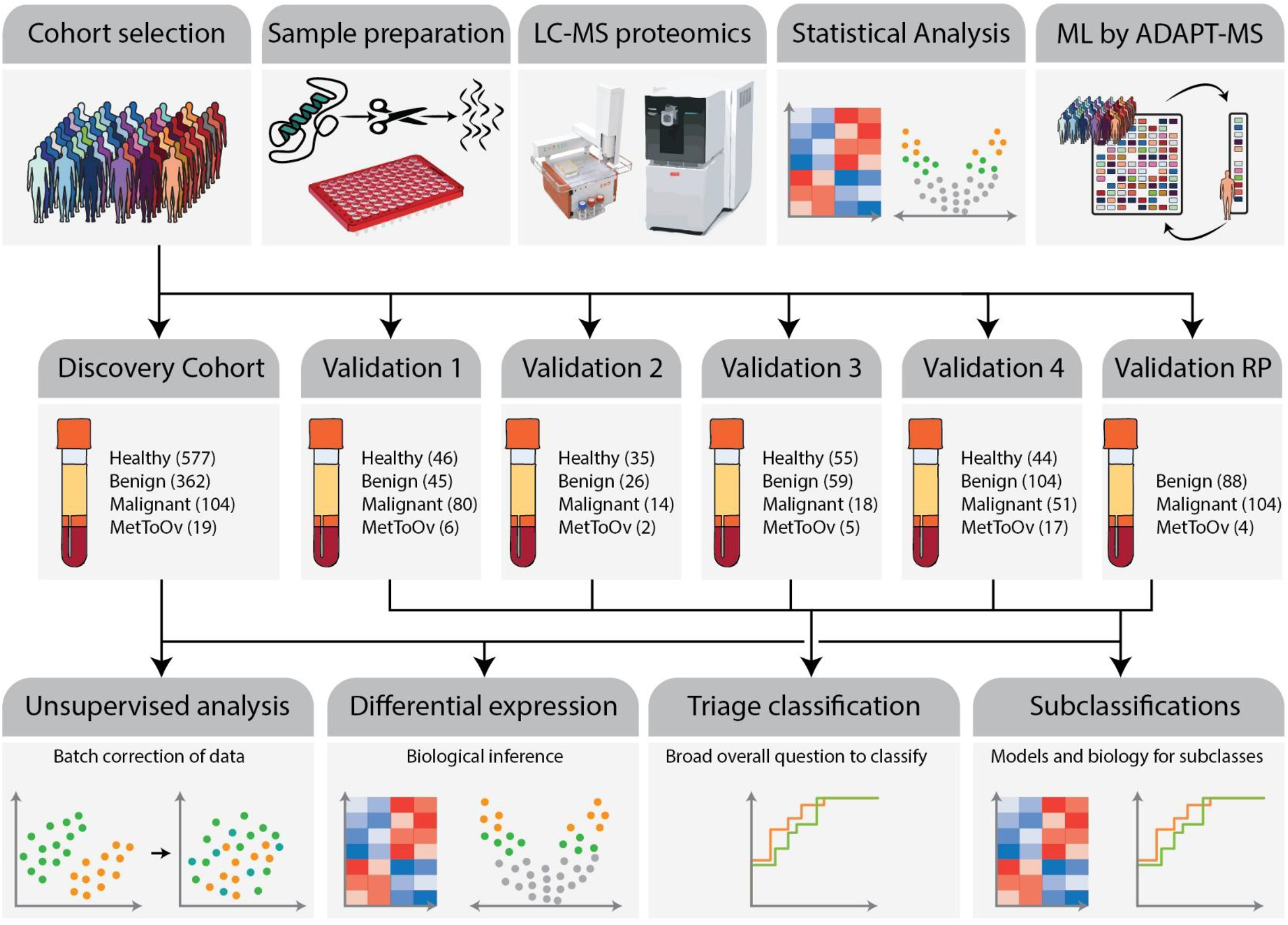
Plasma samples from the discovery cohort (n=1062) and five validation cohorts were processed with the PCA-N high-throughput workflow and analyzed by DIA mass spectrometry on an Orbitrap Astral platform. Proteomic data supported unsupervised analysis, differential-expression analysis, ADAPT-MS machine learning (ML) classification of malignant versus benign adnexal masses, and biological-subgroup characterization; the external Roswell Park (RP) cohort comprises adnexal masses only (no healthy controls). The benign-versus-malignant classification used the benign + malignant subset of each validation cohort (Val1 n=131, Val2 n=42, Val3 n=82, Val4 n=172, ValRP n=196 classifiable). Groups: healthy control (Healthy), benign adnexal mass (Benign), malignant adnexal mass (Malignant), metastasis to ovary (MetToOv).

Table 1 summarizes baseline clinico-pathologic characteristics. The discovery cohort included 1062 women (577 healthy controls, 362 benign adnexal masses, 104 malignant or borderline masses, and 19 MetToOv); the binary discovery training set therefore comprised 485 samples (362 benign, 123 malignant). Four internal validation cohorts (Val1, Val2, Val3, Val4) and one external validation cohort from Roswell Park, (ValRP) were analyzed, representing 131, 42, 82, 172, and 196 classifiable samples, respectively (623 in total). The following category distributions were noted: malignant adnexal mass, MetToOv, benign adnexal mass, and healthy control. Across all cohorts except ValRP, median age was higher in the malignant groups (60 years) than in healthy controls (45 years; p<0.01) or benign masses (44.5 years; p<0.01, Mann-Whitney), consistent with the epidemiology of ovarian cancer, whereas healthy controls and benign masses did not differ in age (p=0.29). Body mass index was modestly lower in the malignant groups (median 26.3 kg/m^2^) than in healthy controls (29.0; p<0.01) or benign masses (28.6; p<0.01) and did not differ between healthy controls and benign masses (p=0.43). Age and body mass index were not recorded for the external Roswell Park cohort; its comparability rests on diagnostic composition and on the concordance of its protein identifications with those of the other cohorts.

**Table 1.**
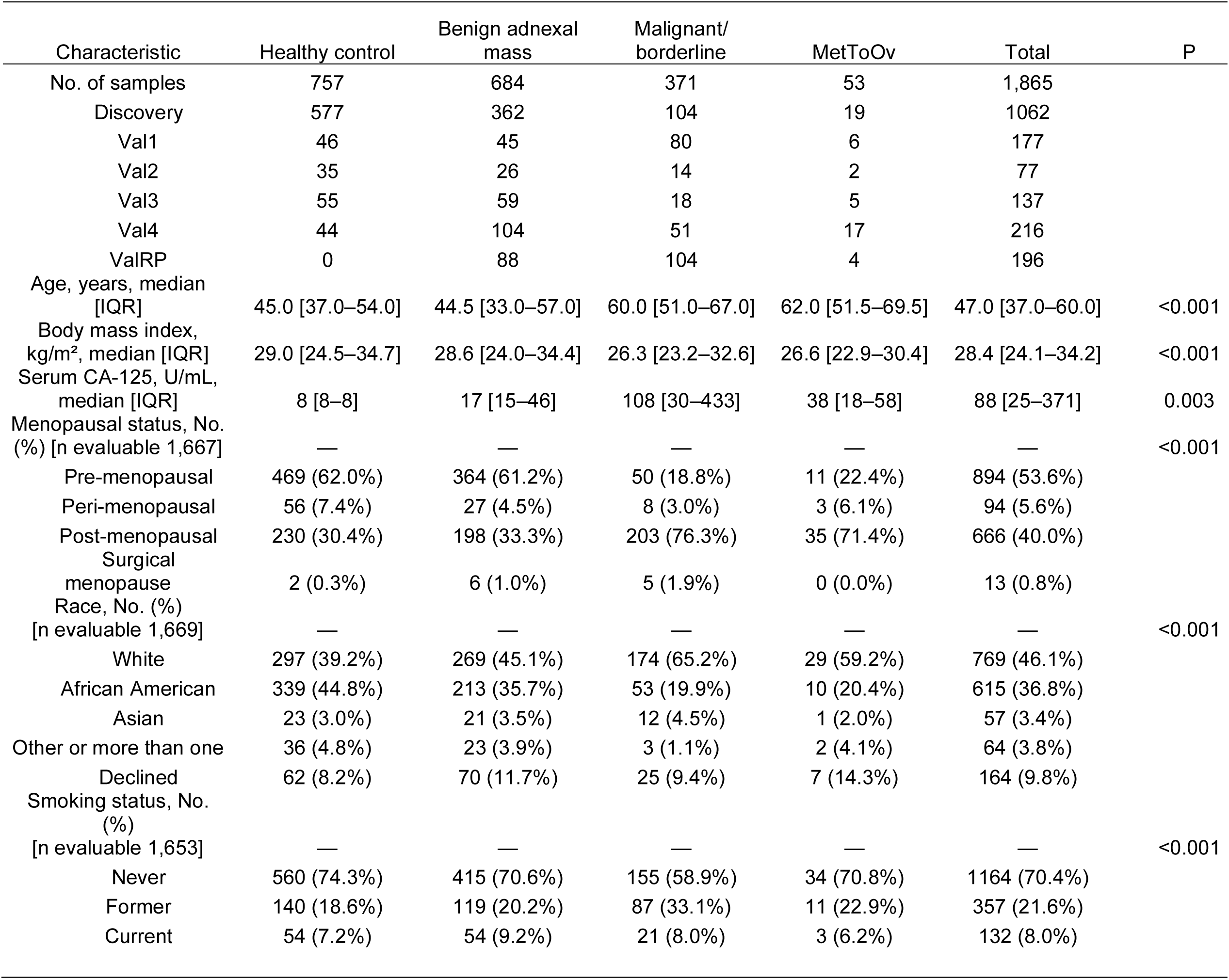
Baseline characteristics of the study population, pooled across the discovery and five validation cohorts. Continuous variables are median [interquartile range] compared by Kruskal-Wallis test; categorical variables are number (percentage of samples with the variable recorded) compared by chi-square test. Percentages are computed within each column over the samples for which the variable was recorded, so denominators differ between variables; the evaluable n is given in each variable heading. Groups: healthy control (Healthy), benign adnexal mass (Benign), malignant adnexal mass (Malignant), metastasis to ovary (MetToOv). Cohorts: University of Chicago Medical Center discovery cohort (2019 – 2022), internal validation (Val) cohorts, Val1, Val2, Val3 and Val4 (2022 – 2025) and at Roswell Park Comprehensive Cancer Center, Buffalo, NY (external validation cohort, ValRP)

| Characteristic | Healthy control | Benign adnexal mass | Malignant/<br>borderline | MetToOv | Total | P |
| --- | --- | --- | --- | --- | --- | --- |
| No. of samples | 757 | 684 | 371 | 53 | 1,865 |  |
| Discovery | 577 | 362 | 104 | 19 | 1062 |  |
| Val1 | 46 | 45 | 80 | 6 | 177 |  |
| Val2 | 35 | 26 | 14 | 2 | 77 |  |
| Val3 | 55 | 59 | 18 | 5 | 137 |  |
| Val4 | 44 | 104 | 51 | 17 | 216 |  |
| ValRP | 0 | 88 | 104 | 4 | 196 |  |
| Age, years, median [IQR] | 45.0 [37.0–54.0] | 44.5 [33.0–57.0] | 60.0 [51.0–67.0] | 62.0 [51.5–69.5] | 47.0 [37.0–60.0] | <0.001 |
| Body mass index, kg/m<sup>2</sup>, median [IQR] | 29.0 [24.5–34.7] | 28.6 [24.0–34.4] | 26.3 [23.2–32.6] | 26.6 [22.9–30.4] | 28.4 [24.1–34.2] | <0.001 |
| Serum CA-125, U/mL, median [IQR] | 8 [8–8] | 17 [15–46] | 108 [30–433] | 38 [18–58] | 88 [25–371] | 0.003 |
| Menopausal status, No. (%) [n evaluable 1,667] | — | — | — | — | — | <0.001 |
| Pre-menopausal | 469 (62.0%) | 364 (61.2%) | 50 (18.8%) | 11 (22.4%) | 894 (53.6%) |  |
| Peri-menopausal | 56 (7.4%) | 27 (4.5%) | 8 (3.0%) | 3 (6.1%) | 94 (5.6%) |  |
| Post-menopausal | 230 (30.4%) | 198 (33.3%) | 203 (76.3%) | 35 (71.4%) | 666 (40.0%) |  |
| Surgical menopause | 2 (0.3%) | 6 (1.0%) | 5 (1.9%) | 0 (0.0%) | 13 (0.8%) |  |
| Race, No. (%) [n evaluable 1,669] | — | — | — | — | — | <0.001 |
| White | 297 (39.2%) | 269 (45.1%) | 174 (65.2%) | 29 (59.2%) | 769 (46.1%) |  |
| African American | 339 (44.8%) | 213 (35.7%) | 53 (19.9%) | 10 (20.4%) | 615 (36.8%) |  |
| Asian | 23 (3.0%) | 21 (3.5%) | 12 (4.5%) | 1 (2.0%) | 57 (3.4%) |  |
| Other or more than one | 36 (4.8%) | 23 (3.9%) | 3 (1.1%) | 2 (4.1%) | 64 (3.8%) |  |
| Declined | 62 (8.2%) | 70 (11.7%) | 25 (9.4%) | 7 (14.3%) | 164 (9.8%) |  |
| Smoking status, No. (%) [n evaluable 1,653] | — | — | — | — | — | <0.001 |
| Never | 560 (74.3%) | 415 (70.6%) | 155 (58.9%) | 34 (70.8%) | 1164 (70.4%) |  |
| Former | 140 (18.6%) | 119 (20.2%) | 87 (33.1%) | 11 (22.9%) | 357 (21.6%) |  |
| Current | 54 (7.2%) | 54 (9.2%) | 21 (8.0%) | 3 (6.2%) | 132 (8.0%) |  |

### Plasma proteome profiling

After quality filtering, we quantified 2076 protein groups and a median of 1119 protein groups per discovery sample by DIA-MS (4% data completeness at protein level), spanning approximately four orders of magnitude in abundance—from abundant complement and coagulation factors to lower-abundance signaling and tissue-leakage proteins. Principal-component analysis confirmed that ComBat correction removed the acquisition-batch structure while preserving expected separation by preprocessing influences and inflammation biology (Supplementary Figure 1, 2).

### Differential protein expression in the discovery cohort

One-way ANOVA across the four diagnostic categories identified 590 proteins with significantly altered abundance in the discovery cohort (q<0.01; Figure 2A). Hierarchical clustering into five clusters revealed category-associated expression patterns, several of which showed a graded change from Healthy through Benign to Malignant and MetToOv, consistent with a proteomic gradient that tracks disease severity. Gene Ontology overrepresentation analysis showed that the clusters were enriched for distinct biological pathways—including complement and coagulation, extracellular matrix, and innate immune programs (Supplementary Figure 3). Pairwise comparison identified 466 proteins differing between benign and malignant adnexal masses (Benign vs Malignant; q<0.05, 335 at q<0.01) and 528 between benign masses and metastasis to ovary (Benign vs MetToOv; q<0.05, 382 at q<0.01; Figure 2B, C). Fold changes for the two comparisons were correlated (Pearson r = 0.695), indicating a shared core of dysregulated proteins in the malignant samples (Figure 2D). Overall, the sets of significantly altered proteins (benign versus malignant / MetToOv) overlap, but the effect sizes are larger in the MetToOv samples. This indicates a shared biology among malignant ovarian tumors and suggests that many circulating biomarkers originate from the ovarian tumor microenvironment in response to the presence of cancer, rather than from a specific histology.

**Figure 2.**
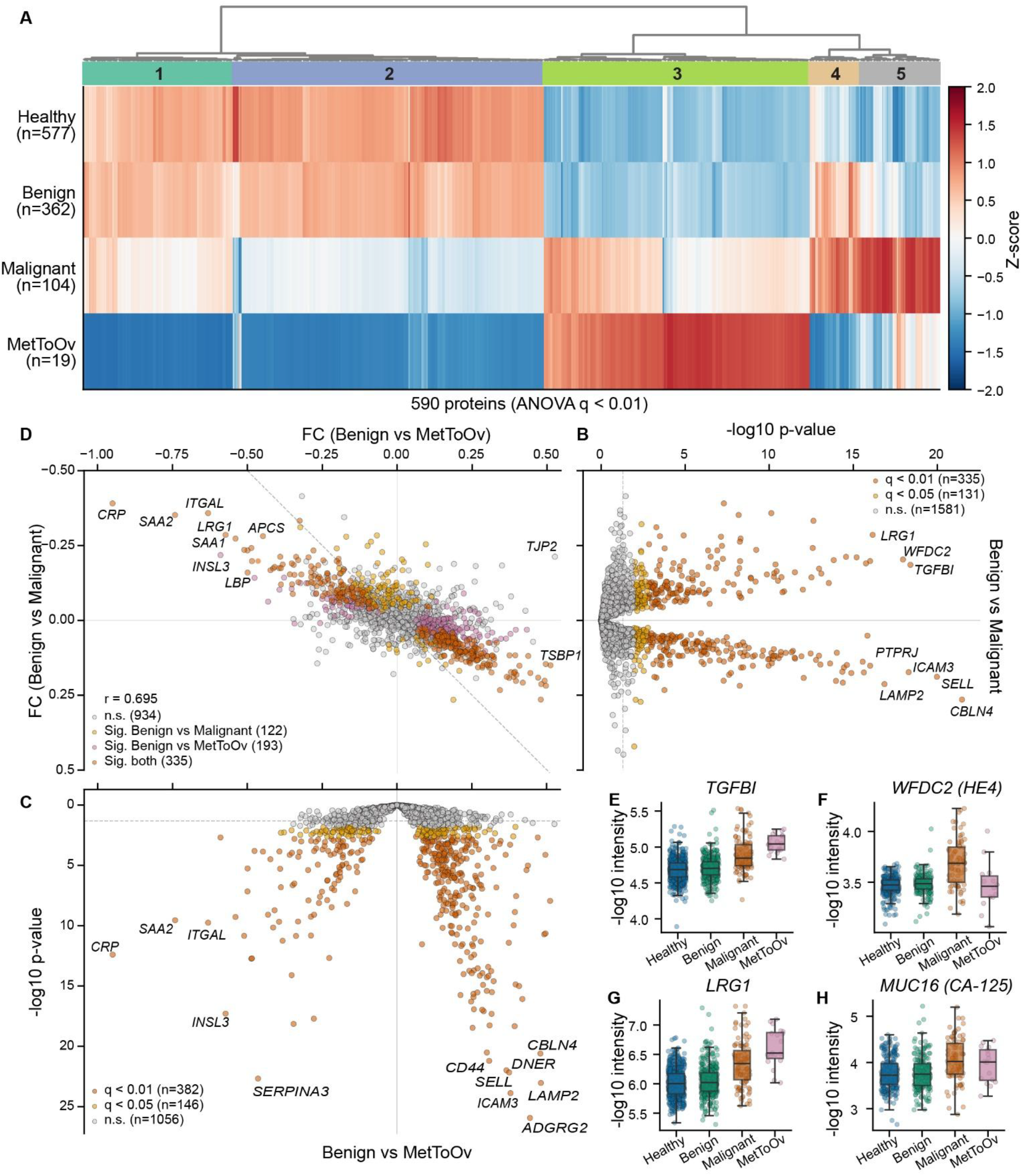
Discovery-cohort plasma proteomics identifies 590 proteins differentially abundant across diagnostic categories. Discovery cohort, n=1,062 (577 healthy controls, 362 benign adnexal masses, 104 malignant/borderline adnexal masses, 19 metastases to ovary); log10 intensities, batch-corrected, not imputed. (A) Hierarchical clustering heatmap of proteins significant by one-way ANOVA across the four categories (q<0.01; 591 of 2,076 tested, the 590 proteins with a mean in every category are shown). Color is the Z-scored category mean; the bar above marks the five clusters (enrichment in Supplementary Figure 3). (B) Volcano plot, benign versus malignant adnexal mass: 335 proteins at q<0.01, 466 at q<0.05. (C) Volcano plot, benign adnexal mass versus metastasis to ovary: 382 at q<0.01, 528 at q<0.05. (D) Fold changes of the two comparisons, benign-versus-malignant on the x-axis and benign-versus-metastasis on the y-axis (Pearson r=0.695), colored by significance in one, the other or both comparisons. (E–H) Abundance of TGFBI (E), WFDC2/HE4 (F), LRG1 (G), and MUC16/CA-125 (H) across the four categories; boxes give median and interquartile range with all samples overlaid. Significance colors throughout all figures: q<0.01 vermillion, q<0.05 orange, not significant grey.

Among individual proteins, TGFBI (transforming growth factor-β-induced protein) showed the largest benign-versus-malignant fold change, and WFDC2 (HE4), an established ovarian cancer biomarker, was significant only in the benign-versus-malignant comparison but not in benign-versus-MetToOv. LRG1, a marker of inflammation and angiogenesis (Wu et al. 2015), and several complement factors (C3, C4A, CFB) (Ricciuti et al. 2025) were elevated in the malignant groups (Figure 2E–H).

### Metadata structure and confounders affecting the proteomic signature associated with a malignant adnexal mass

A subtype comparison confirmed that the malignant signature is driven predominantly by epithelial tumors (Supplementary Figure 4 A, B). The three most significantly upregulated proteins in malignant epithelial tumors compared to benign epithelial masses were TGFBI, SERPINF2, and SERPINA1. CA-125 and HE4 rank 62nd and 19^th^, respectively, among 225 proteins significantly upregulated (q<0.05) in malignant epithelial tumors. When performing a cross-comparison of benign-to-malignant differences in epithelial and non-epithelial tumors, we found minimal overlap in the altered protein signatures, reflecting the distinct biology of tumors of different embryologic origins (Supplementary Figure 4C). CD276 was the only protein significantly altered in non-epithelial malignant tumors vs benign adnexal masses (Supplementary Figure 4D).

To explore how clinical and demographic variables shape the plasma proteome and whether they confound the malignancy signal, we mapped the discovery proteome onto the main metadata parameters. Proteins significantly associated with sample category, age, race, menopausal status, or BMI formed a co-expression network of 505 connected proteins that partitioned into four modules, two dominated by the diagnostic (sample-category) axis and one each by race and by mixed demographic drivers (Supplementary Figure 4E). We then tested whether the benign-versus-malignant protein signal was confounded by the four dominant covariates (age, race, BMI, menopausal status). For each of the 553 proteins that were significantly differentially abundant between benign and malignant masses, variance was partitioned among malignancy and the covariates using multivariable ANOVA to ascertain which proteins were predictive irrespective of covariates. Malignancy remained significant after adjusting for age, BMI, race, and menopausal status in 523 of 553 proteins (95%), indicating a signal largely independent of these variables (Supplementary Figure 4F); however, for 167 proteins (30%) a covariate—most often race (n=93) or BMI (n=59)—explained more variance in the proteomic results than a diagnosis of malignancy (Supplementary Figure 4G). Overall, BMI was the dominant covariate accounting for changes in protein with the largest covariate partial η^2^ (mean of 0.026) and most proteins influenced by it (n=198). Several proteins (e.g. LUM, CPN2, VCAM1) were significantly different between the benign and malignant groups and showed distinct trajectories across different confounders. Lumican (LUM), a class II small leucine-rich extracellular-matrix proteoglycan with context-dependent tumour-promoting and tumour-suppressive activity in tumour stroma, illustrates this (Wolff et al. 2019; Chen et al. 2020). In benign samples LUM increased with BMI, whereas in malignant samples it did not, producing a significant BMI × diagnosis interaction and a benign– malignant separation that was largest in the normal-BMI range and absent above BMI ≈ 40 (Supplementary Figure 4H). At matched BMI, LUM was higher in self-reported Black women than White women. Consequently, the discriminative value of LUM was BMI-dependent, as is the case, for CPN2 and VCAM1, where an age dependency is observed (Supplementary Figure 4I–J).

### ADAPT-MS classification performance

We recently described Adaptive Diagnostic Architecture for Personalized Testing by Mass Spectrometry (ADAPT-MS), which enables the direct use of discovery-mode proteomics data for diagnostic and prognostic interpretation at the level of individual samples (Müller-Reif et al. 2026). Briefly, prior information is required in the form of one or more discovery cohorts that are used two-fold: a) to extract proteins by t-test that are different between benign and malignant individuals in the discovery cohort; and b) after filtering the relaxed feature list (from a)) for proteins detected in a sample that undergoes classification, to train a sample-specific classifier for that sample (Figure 3A).

**Figure 3.**
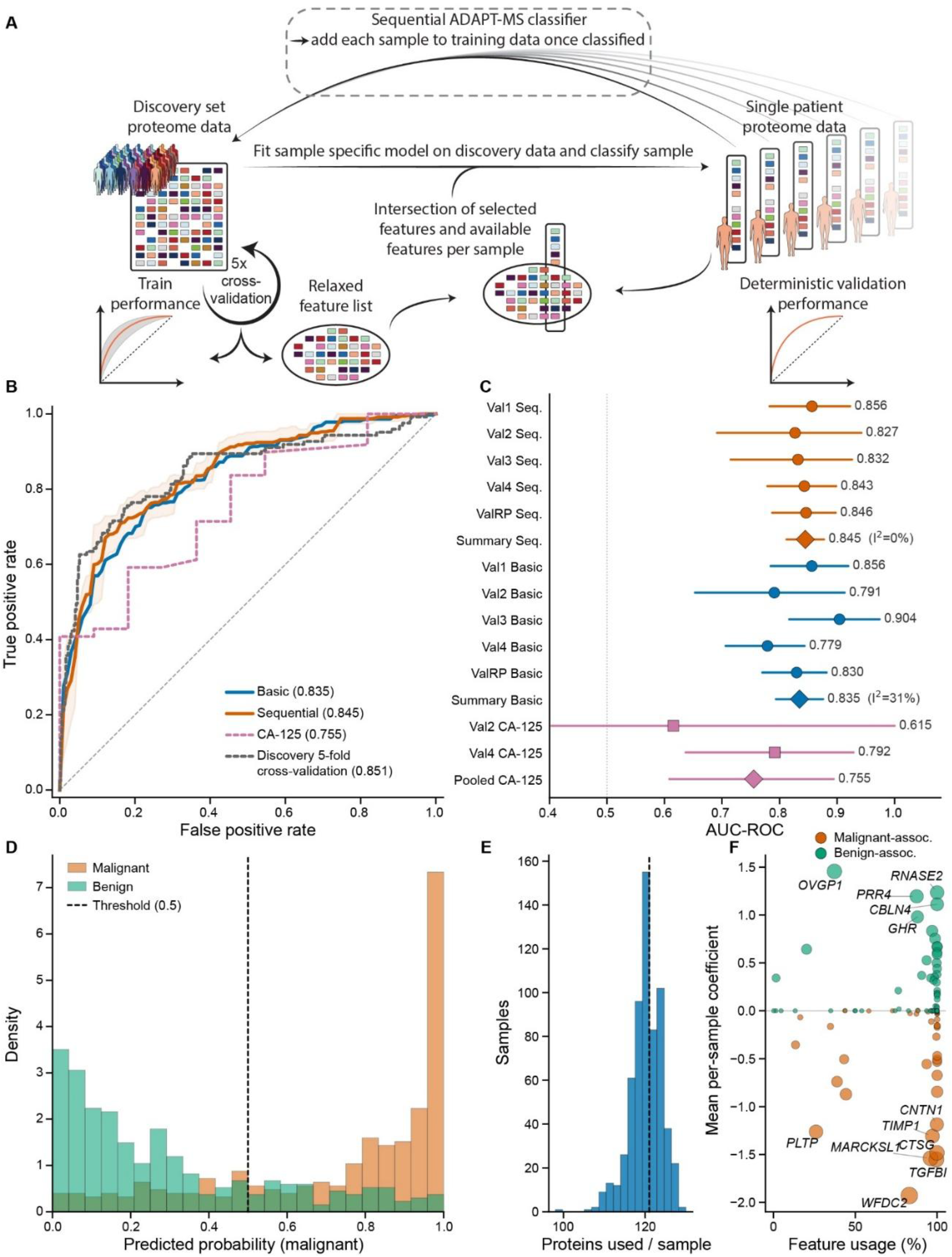
The sequential ADAPT-MS classifier discriminates malignant from benign adnexal masses across five validation cohorts. Validation samples, n=623 (Val1 131, Val2 42, Val3 82, Val4 172, external Roswell Park 196); scores are predicted probability of malignancy, threshold 0.5. (A) Schematic of the ADAPT-MS architecture for the basic and sequential configurations. (B) ROC curves as the mean across the five cohorts for the basic and sequential classifiers, with ±1 SD across cohorts shaded for the sequential classifier. Legend values are random-effects summary AUCs for the two configurations and for held-out discovery (grey dashed, five-fold cross-validation), and the pooled AUC for clinical serum CA-125 on its matched subset. (C) Forest plot of point AUC with 95% bootstrap CIs per cohort. Diamond giving the random-effects summary with I^2^. (D) Density-normalized histograms of predicted malignancy probability for malignant and benign samples, pooled across cohorts, with the 0.5 threshold marked (>=0.5 is classified as malignant). (E) Number of proteins selected for the classifier per validation sample (median 121 of the 138 used anywhere). (F) Summary statistics of feature (protein) usage against their mean per-sample ElasticNet coefficient; malignancy-associated proteins are in orange and benign-associated proteins in green, point size ∝absolute coefficient; 36 discovery-selected features never identified in validation are excluded.

Trained on the 485-sample binary discovery set and validated across the five cohorts, the basic ADAPT-MS classifier achieved a random-effects summary AUC of 0.835 (95% CI, 0.794–0.876), with modest between-cohort heterogeneity (I^2^=31%; range, 0.779–0.904). Compared with training performance obtained from the discovery cohort in a five-fold cross-validation manner, which gave a random-effects summary AUC of 0.851 (95% CI, 0.805–0.896), validation performance was not significantly different (difference +0.006; 95% CI, −0.050 to +0.062; p=0.836).

The cohort structure in the current study allows us to apply our recently proposed adaptive learning strategy with ADAPT-MS. To this end, we built a sequential classifier that adds each sample from the validation cohorts to the discovery set after it has been classified. The sequential ADAPT-MS classifier achieved cohort-specific AUCs of 0.856, 0.827, 0.832, 0.843 and 0.846, giving a random-effects summary AUC of 0.845 (95% CI, 0.812–0.877). Between-cohort heterogeneity was undetectable (I^2^=0%), indicating consistent discrimination across institutions, including the external Roswell Park cohort (0.846) (Figure 3B). The full comparison of ADAPT-MS configurations shows that the sequential classifier performs more consistently and, on average, better than the basic classifier in all but validation cohort #3 (Figure 3C, Supplementary Table 2). Briefly, the classifier calculates a probability score between 0 and 1 for each sample, with a default cutoff of 0.5: Samples with a score >=0.5 are classified as malignant and otherwise as benign. The distribution of those probability scores helps interpret the classifier’s performance and balance, and shows a clear separation of malignant from benign samples with apparent signal present for both sample classes (Figure 3D).

Feature-importance analysis identified WFDC2 (HE4), TGFBI, MARCKSL1, and CTSG as among the highest-weighted proteins, and each per-sample model drew on a median of 121 proteins (of 138 detected across the validation cohorts) (Figure 3E). This illustrates the panel-free adaptivity of the framework. The strength of the model is that it does not rely on a complete dataset; instead, it adapts to the proteins actually detected in each sample and then fits the optimal classifier for those proteins on the training set (Figure 3F). Indeed, sequential ADAPT-MS has a very balanced sensitivity and specificity, whereas the basic model performs better in terms of sensitivity (0.904) but has very limited specificity (0.435) (Supplementary Table 2). Finally, discrimination remained stable across the number of features selected per split (5–200 features), confirming that performance is not sensitive to this design choice. The sequential classifier including the entire dataset (with the discovery) performed very well with 125 proteins – adding more proteins did not improve the performance (Supplementary Figure 5). The top proteins driving the difference between benign and malignant masses include HE4 (WFDC2), TGFBI and MARCKSL1, which shift the prediction toward malignancy with mean per-sample coefficients of −1.93, −1.55 and −1.54 and are selected in 83.0%, 99.5% and 96.0% of validation samples, respectively. The strongest features associated with a benign adnexal mass are OVGP1 (+1.46, selected in 37.2% of samples), RNASE2 (+1.24, 100%) and PRR4 (+1.20, 87.5%). HE4 is the single most influential of the 138 features the classifier uses, whereas CA-125 (MUC16) ranks 28^th^ overall and 15^th^ among the 46 malignancy-associated features, with a mean coefficient of −0.51 and selection in 43.2% of samples — roughly half the usage and a quarter of the weight of HE4 (Figure 3F).

In a cross-analysis of prediction results versus potentially confounding factors we find no relevant correlation of age, BMI, menopausal status or race with the outcome if a sample was predicted benign or malignant (Supplementary Figure 6). Further we expanded this analysis to technical factors like proteome depth of the measurement per sample and plasma pre-processing biases like erythrocyte, platelet, PBMC and coagulation contamination, where no relevant influence of those was observable on the prediction outcomes (Supplementary Figure 7).

### Integration with O-RADS ultrasound and serum biomarkers

Pre-operative O-RADS ultrasound and serum CA-125 were available for only a subset of patients, and their availability varied markedly across cohorts: O-RADS scores were present for 47 of 131 Val1, 20 of 42 Val2, 48 of 82 Val3, and 116 of 172 Val4 patients but were unavailable for the Roswell Park cohort, and serum CA-125 measured by ELISA was available for 125 patients across Val1, Val2, Val3, and Val4 (Supplementary Figure 10A). The proteomic feature panel was available for every patient.

Among patients with pre-operative O-RADS (n=231; Val1, Val2, Val3, and Val4), the ADAPT-MS proteomic classifier discriminated malignant from benign masses with an AUC of 0.852, comparable to O-RADS alone (0.863). Adding the O-RADS score to the proteomic model increased discrimination to 0.922, the highest observed in the study (Figure 4A). The per-sample probability shifts (Figure 4B) showed that O-RADS primarily corrected a subset of proteomically ambiguous cases rather than uniformly rescaling predictions. Five benign and five malignant cases correctly classified by proteomics were falsely classified by the proteomics + O-RADS classifier, whereas ten benign and seven malignant cases were correctly reclassified by adding the O-RADS classifier (Figure 4B, n=231, threshold 0.5). Adding proteomics to O-RADS resolves 16 malignant cases missed by ultrasound alone and three misclassified benign cases, at the cost of four newly misclassified malignant and seven benign cases, resulting in a net gain of 12 malignant cases against a net loss of four benign cases (Figure 4C). Against serum CA-125, the proteomic classifier was superior on the matched subset (AUC 0.915 vs 0.777; n=125). Proteome-derived surrogates of established assays performed similarly to, but did not exceed, the full proteomic model: a ROMA-like model (proteomic MUC16 and WFDC2 with menopausal status and age (Molina et al. 2011)) achieved an AUC of 0.833 compared to 0.867 for proteomics on the same patients (n=207), and a two-marker model (proteomic MUC16 and WFDC2) reached 0.763 versus 0.890 (n=239; Figure 4D). Individual proteins —proteomic WFDC2 (HE4) or MUC16 (CA-125)—were substantially weaker classifiers than the multi-protein model across all validation cohorts (Supplementary Figure 8). Together, these matched comparisons indicate that the multi-protein classifier captures the discriminative information of single biomarkers and outperforms CA-125 alone, while ultrasound contributes complementary information when combined with proteomics.

**Figure 4.**
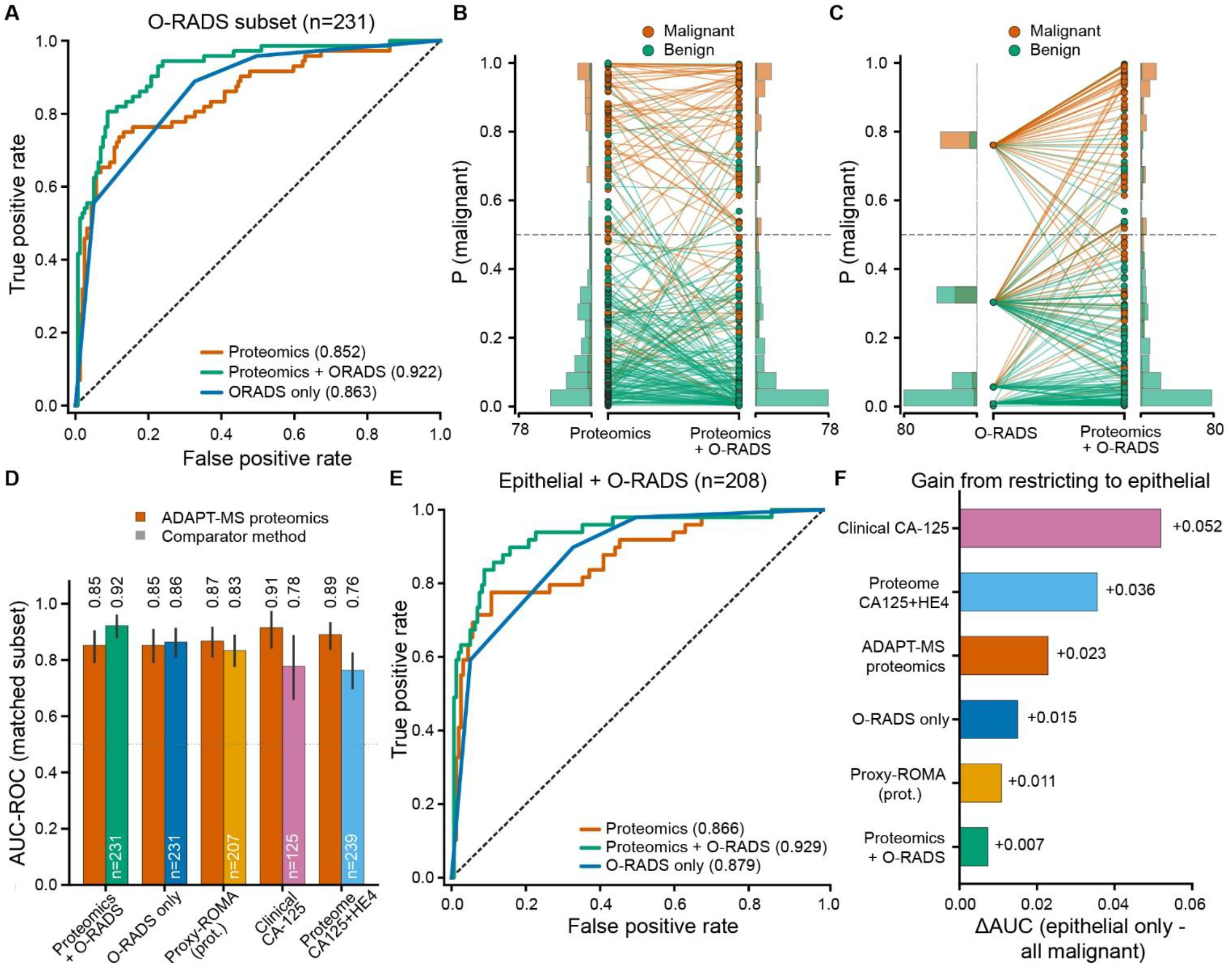
Integration of O-RADS ultrasound and single biomarkers with ADAPT-MS proteomics. (A) ROC curves on the O-RADS subset (Val1–Val4, n=231) for proteomics, O-RADS alone (0.863), and proteomics plus O-RADS. (B) Per-sample predicted probability of malignancy under proteomics versus proteomics plus O-RADS on the same subset; each sample drawn as a line between the two models and colored by true label (vermillion malignant, green benign); marginal histograms show the score distributions and the dashed line the 0.5 threshold. (C) The same comparison for O-RADS versus O-RADS plus proteomics. (D) Matched 1:1 comparison of the proteomic classifier against each comparator, each evaluated on that comparator’s own available subset (n annotated): proteomics plus O-RADS and O-RADS alone, a proteome-derived ROMA-like model, clinical serum CA-125 and a two-marker proteomic model of MUC16 and WFDC2 (n=239). Bars give AUC with 95% bootstrap confidence intervals. (E–F) Epithelial ovarian cancer: malignant cases scored are limited to the epithelial lineage without retraining any model (the external Roswell Park cohort has no histological subcategory and is excluded). (E) ROC curves on the epithelial O-RADS-available subset (Val1–Val4, of which 49 epithelial cancers) for proteomics, O-RADS alone, and proteomics plus O-RADS. (F) Change in AUC from restricting to epithelial malignancy for each method, ordered by gain.

Decision-curve analysis showed a positive net benefit for ADAPT-MS across many clinically relevant thresholds, outperforming clinical strategies in which all patients presenting with an adnexal mass are treated (“treat-all”) or no patient are treated (“treat-none”) (Supplementary Figure 9A). At the Youden-optimal threshold (0.37) sensitivity was 0.850 and specificity was 0.720. Calibration showed good agreement between predicted and observed probabilities (Supplementary Figure 9B, C). On the O-RADS-available subset, the combined proteomics-plus-O-RADS classifier demonstrated a consistent clinical-utility profile, with higher specificity at the balanced operating point (Youden-optimal sensitivity 0.806, specificity 0.912; Supplementary Figure 10). Discrimination remained strong across clinically important subgroups: Among post-menopausal women (n=255), the AUC was 0.824 (95% CI 0.772– 0.872), and in the age-matched 50–70-year subgroup (n=191) it was 0.799 (0.730–0.861), supporting age-independent classification despite the expected age imbalance between benign and malignant groups (Supplementary Figure 11).

When restricting the malignant group to epithelial cancers only, classification performance of the proteomics (AUC 0.866) and the proteomics + ORADS (AUC 0.929) classifier was comparable to the complete cohort (Figure 4E). The classifiers relying purely on CA-125 or proteomics measured MUC16 and HE4 gained most from this restriction to epithelial cancers (Figure 4F).

## DISCUSSION

We show that plasma proteomics, interpreted using the ADAPT-MS framework, distinguishes malignant from benign adnexal masses across five independent cohorts, including an external academic center, with a summary AUC of 0.835, and that it is complementary to routine ultrasound. To our knowledge, this is the largest plasma proteomic study focused on adnexal-mass classification and the first application of an adaptive, panel-free learning framework in diagnostic use or proteomics in general.

This approach differs from existing tools by incorporating data from more than 150 proteins, rather than relying on a few analytes such as CA-125, HE4, and ROMA. This enables the classifier to capture a broader range of biological signals indicative of malignancy. The matched comparisons in Figure 4 make this concrete: among patients with available data, the proteomic classifier exceeded serum CA-125 (AUC: 0.915 vs 0.777) and matched the performance of proteome-derived ROMA-like and two-marker surrogates, indicating that the multi-protein model already contains the discriminative information of the single biomarkers while adding to it. Most importantly for clinical translation, proteomics proved complementary to imaging: the proteomic classifier alone performed comparably to O-RADS, but combining the two produced the strongest discrimination in the study (AUC 0.922). For clinical application, this supports a deployment approach in which proteomics serves as an adjunct to a standardized ultrasound exam—particularly clinically valuable for women with indeterminate O-RADS or ADNEX assessments—rather than as a replacement for imaging.

The ADAPT-MS framework enables direct diagnostic use of discovery-mode proteomics, eliminating the need for targeted-assay development or a locked panel (Müller-Reif et al. 2026). This flexibility is well suited for the clinical challenge of evaluating adnexal masses, where histologic heterogeneity (epithelial, non-epithelial, borderline, and metastatic disease) cannot be represented by a narrow fixed panel optimized for a single histology, and it underlies the availability advantage seen in Figure 4A, where proteomic features were measurable in every patient whereas O-RADS and CA-125 were missing for many. Overall, the sequential learning approach leverages a key characteristic of how women diagnosed with an adnexal mass are treated: patients identified as potentially malignant undergo surgery, and the pathology results provide a confirmed label that can be reused to enhance the model. This establishes a self-improving cycle unique to surgically treated patients in which pathology as the gold standard for diagnosis is available. A limitation in our test setting is that all classified samples, including those not recommended for surgery, are treated as unblinded and continuously added to the training cohort. While the true labels of individuals not undergoing surgery could not be included sequentially, this shows that cohort size does matter (given the variety and composition of each validation set used here), as performance improves with a larger training dataset and the stability of the classifier increases. In theory, specific patient-targeting subcohorts could be further stratified by covariates such as age, menopausal status, race, or BMI that may improve the specificity of the classifier; however, in practice, these approaches reduce the size of the training data (with more subgroups) and thus diminish predictive performance at the population-level. Our results suggest that once a critical mass of samples and data is reached, the growing sample cohort using the ADAPT-MS slicing approach may improve predictive performance, by access to a sufficient size of specific subgroup samples of confounding factors, like age, BMI or race groups.

Our biological analysis indicates that the malignant signature is driven predominantly by epithelial tumors, with 495 significantly altered proteins in the epithelial comparison but only one in the non-epithelial comparison. Combined with ORADS, predictability improved to an AUC = 0.922, however, this component itself only improved predictability minimally. Since non-epithelial tumors account for about 10% of ovarian cancers, the classifier’s effectiveness in detecting epithelial disease covers most cases. However, better classification of non-epithelial tumors will need larger or subtype-specific cohorts (Colombo et al. 2019). While this cohort demonstrates improved prediction, it is important to note that the difference between this histology-specific subgroup and the analysis with all histologies shows only minor changes in predictability and should not be restricted. Therefore, restricting the assay only to epithelial histologies is likely unnecessary, supporting the use of this “mass” proteomic signature across all suspected adnexal masses independent of histology.

Many observed proteomic changes are associated with remodeling of the extracellular matrix and the host local ovarian tissue response as well as systemic adaptations to malignant transformation (Figure 2, Supplementary Figure 3). Indeed, recent single-cell RNA sequencing (scRNA-seq) data have shown specific gene expression changes in the TME, as well as proliferation and reprogramming of specific stromal and immune cell types that reorganize the entire tumor organ (Jiménez-Sánchez et al. 2020; Balkwill et al. 2026). The consistently observed plasma proteomic changes in epithelial invasive tumors likely reflect TME remodeling during invasion by surrounding stromal and immune cells, rather than the cancer cells themselves. Non-epithelial tumors which differ in their embryonic cell of origin show no overlap of protein signature and no correlation in protein fold-changes to the epithelial tumors. Importantly, the protein signature predicted malignancy independently of common confounders reported with other serum markers, showing no difference in predictive performance across menopause status, BMI, and age. Early FIGO stage had a lower prediction index (AUC=0.712) compared to FIGO stages II-IV (AUC II 0.851, III 0.898, IV 0.896), which may reflect a more contained biologic response to malignancy and fewer proteins secreted into blood.

This study has several limitations. First, it was conducted at two academic referral centers with a relatively high prevalence of ovarian cancer; performance should therefore be validated in broader clinical settings, particularly in lower-prevalence community populations. Because ultrasound quality was high in our cohort, the relative value of a proteomics-based classifier may be even greater where expert ultrasound is less consistently available.

Second, the primary sequential classifier assumes that pathology-confirmed outcomes are available for all previous cases before scoring the next case. In practice, this assumption will not fully hold: patients predicted to have malignancy are typically operated on promptly, whereas those predicted to have benign disease are less likely to have immediate pathological confirmation. Real-world performance is therefore expected to fall between that of the static Basic classifier and the fully label-informed sequential model. A practical initial implementation would include routine follow-up imaging for patients classified as benign, both to support standard clinical care and to enable outcome auditing.

Third, comparisons with O-RADS and CA-125 were limited to subsets with available data, which differed in size and case mix. HE4 was not routinely measured at either institution, precluding direct comparison with the clinical ROMA algorithm; similarly, direct comparison with ADNEX was not possible in a fully matched dataset. The proteome-derived ROMA-like surrogate used here should be regarded only as an approximation of the clinical assay.

Fourth, some clinically relevant subgroups, including non-epithelial malignancies and BRCA carriers, were small and require further study. Fifth, although classifier discrimination was robust across age and menopausal status, the age imbalance between benign and malignant groups reflects the underlying epidemiology of the disease and remains an important consideration in interpretation.

Future studies should prospectively validate this approach in community settings and include concurrent assessment with CA-125, HE4, and standardized ultrasound models such as O-RADS and ADNEX. The proteomics-plus-ultrasound strategy should also be formally tested, and automated clinical reporting tools developed to support implementation. As population-scale proteomics datasets continue to grow, this framework should become increasingly refined over time.

A single plasma proteomic measurement interpreted with ADAPT-MS classified adnexal masses with consistent performance across institutions, matched O-RADS, outperformed CA-125 alone, and performed best when combined with ultrasound. By operating directly on discovery-mode mass spectrometry data, this approach provides a practical framework for proteomics-guided surgical decision-making in women with adnexal masses and improved current fixed biomarker strategies.

## PATIENTS AND METHODS

### Study design and participants

This multicenter observational study (Figure 1) consecutively enrolled women presenting with adnexal masses at the University of Chicago Medical Center (discovery cohort (2019 – 2022) and internal validation (Val) cohorts, Val1, Val2, Val3 and Val4 (2022 – 2025)) and at Roswell Park Comprehensive Cancer Center, Buffalo, NY (external validation cohort, ValRP). Both institutions are large urban academic centers where multispecialty care is provided through robust gynecology and gynecologic oncology programs, with pathology review by expert gynecologic pathology-trained pathologists. The study was approved by the institutional review boards of both institutions, and all participants provided written informed consent. Reporting adheres to the STARD guidelines for diagnostic accuracy studies and the TRIPOD guidelines for prediction model studies (Bossuyt et al. 2015; Collins et al. 2024).

Women were eligible to enroll if they presented with an adnexal mass documented on imaging (ultrasound, CT, or MRI) and were undergoing planned surgical evaluation. Women who opted for observation of an adnexal mass were also included to represent a practical clinical population and were considered to have a benign mass if 3 consecutive ultrasounds and CA-125 measurements showed changes of less than 10%. Healthy controls included women undergoing benign gynecological surgery (for example, hysterectomy for fibroids or urogynecologic reasons) and were confirmed to be without adnexal pathology. On final review, exclusion criteria included women receiving active chemotherapy at the time of blood collection, a prior malignancy within the past five years, and technical exclusions, such as insufficient sample volume or disrupted processing.

Participants were assigned to four categories depending on the surgical pathology: healthy control (Healthy), Benign adnexal mass (Benign), malignant or borderline adnexal mass (Malignant), and metastasis to ovary (MetToOv). For classification, Malignant and MetToOv were pooled as malignant but Benign did not include the healthy controls. This binary framework reflects the operative clinical question—whether a woman requires referral to a gynecologic oncologist for expert cancer surgery because of a high suspicion for cancer. Benign adnexal masses included epithelial and non-epithelial masses. Malignant adnexal masses included both primary ovarian malignancies and borderline malignancies (i.e., masses that require surgery). Metastasis to the ovary (MetToOv) refers to secondary malignancy of the ovary or other primary malignancies that have spread to the ovaries enlarging the ovary, including gastrointestinal, breast, and other gynecologic malignancies. See Table 1 and Supplementary Table 1 for more details.

The study was approved by the Institutional Review Board at The University of Chicago and Roswell Park Cancer Institute, and informed written consent was obtained from all women participating in the study.

### Sample collection and preparation

Peripheral blood was collected at the University of Chicago or Roswell Park in EDTA tubes and processed to plasma within two hours. All plasma samples were collected using a strict sample collection and processing protocol including 2 rounds of centrifugation as set by the NCI Biorepositories and Biospecimen Research Branch’s best practices for biospecimen collection (NCI 2026 (4th Edition), 2020). Aliquots were stored at −80°C until analysis. Plasma proteomics sample preparation was performed with the PCA-N (perchloric acid precipitation with neutralization) high-throughput workflow, which enables reproducible deep plasma profiling at scale (Albrecht et al. 2025; Viode et al. 2023). In brief, plasma (5 μL) was diluted in 20 μL ddH_2_O, followed by the addition of 25 μL 1 M perchloric acid. Samples were agitated at 4 °C for 1 h, followed by centrifugation at 4000 × g for 20 min at 4 °C. The supernatant (24 μL) was collected and combined with 8 μL 1.4 M sodium hydroxide solution to adjust pH to 8–8.5. Lysis buffer (8 μL; containing 40 mM chloroacetamide, 20 mM DTT, 0.01% DDM, 60 mM TEAB) was added. Proteins were digested using trypsin/LysC (1.6 µL: 0.4 µL trypsin [0.5 µg/µL] 0.4 µL LysC [0.5 µg/µL], 10.5 µL 1 M TEAB, 0.7 µL water), and digestion was stopped with trifluoroacetic acid (TFA, final concentration 0.5%). The peptide mixtures were analyzed by LC-MS/MS.

### Mass spectrometry analysis

Peptides were separated on an Evosep One system (Evosep) (Bache et al. 2018) coupled to an Orbitrap Astral mass spectrometer (Stewart et al. 2024; Hendricks et al. 2024) (Thermo Fisher Scientific). For each sample, 200 ng of peptides were loaded onto C-18 tips (Evotip Pure, Evosep) according to the manufacturer’s protocol. Chromatographic separation was performed on an 8 cm Aurora Rapid XT UHPLC column (AUR3-80150C18-XT, Ionopticks) at 50 °C using the “100 samples per day” method with pre-formed gradients and a total runtime of 11.5 min per sample. Mobile phases consisted of 0.1% formic acid in water (buffer A) and 0.1% formic acid in acetonitrile (buffer B).

Mass spectrometric analysis was performed using a data-independent acquisition (DIA) method (Guzman et al. 2024). The ion source was operated with a static spray voltage of 1900 V in positive ion mode, and the ion transfer tube temperature was set to 280 °C. FAIMS (high-field asymmetric waveform ion mobility spectrometry) was utilized in standard resolution mode with a compensation voltage (CV) of −40 V and a carrier gas flow of 3.5 L/min. Full MS1 scans were acquired in the Orbitrap analyzer at a resolution of 120,000 FWHM over a scan range of 380–980 m/z. The RF lens was set to 40%, and a normalized AGC target of 500% was used, with a maximum injection time of 3 ms. DIA MS2 scans covered the same mass range (380–980 m/z) divided into 150 isolation windows of 4 Th each with window placement optimization enabled. MS2 spectra were acquired with an HCD collision energy of 25%, a normalized AGC target of 500%, and a maximum injection time of 7 ms. The scan range for fragment ions was set to 150– 2000 m/z. Data were collected in profile mode for MS1 and centroid mode for MS2 scans. The expected chromatographic peak width was set to 5 s, and advanced peak determination was enabled to optimize the duty cycle.

### MS raw data processing for discovery cohort

Each raw file was converted to the mzML format using MSConvert. The resulting mzML files were processed using DIA-NN (version 1.8.1) (Demichev et al. 2020) on a high-performance computing cluster. The searches were performed against a human UniProt Swiss-Prot isoform database (downloaded 06/2023) that included oxidation and N-terminal acetylation modifications using the DIA-NN built-in in silico library prediction. All files were analyzed with match-between-runs enabled using the “--use-quant” and “--reanalyse” parameters. The following parameters were applied: peak centering, smart profiling, retention time profiling, and relaxed protein inference. Further, mass accuracy was set to 10 ppm for both MS1 and MS2 scans and the scan window to 7. False discovery rate was controlled at 1% at the peptide-to-spectrum match level.

### MS raw data processing for validation cohorts

Each raw file was converted to the mzML format using MSConvert. The resulting mzML files were processed using DIA-NN (version 1.8.1) on a high-performance computing cluster. The searches were performed against the spectral library generated from the discovery cohort search. Search results in form of pg_matrix.tsv from each validation cohort raw file against the discovery cohort spectral library were used for machine-learning classification on a per-sample basis.

### Data processing

For the discovery cohort, we acquired 1334 LC-MS/MS runs. Of these, 32 were quality-control and reference injections, while 102 were blank runs. Of the 1200 patient-assigned runs, 118 were excluded by study design: 61 plasma samples taken before interval surgeries following neoadjuvant chemotherapy, 16 benign inflammatory conditions, 13 minimal residual disease or chemo response monitoring samples, 20 samples of the subcategory “malignant gynecologic cancer without ovarian involvement”, and 8 samples flagged as not evaluable. Of the resulting 1082 samples, 20 did not pass quality control because of low protein identification counts (below 750 protein identifications; 1.8% (20/1082).

For the final 1062 discovery samples, proteins identified in less than 4% of samples were removed and protein-group intensities were log10-transformed. Missing values in the discovery training matrix were imputed by k-nearest-neighbor imputation (k=5), and acquisition-plate batch effects were corrected with ComBat (Johnson, Li, and Rabinovic 2007). Missing values from the original data were removed after batch correction for any statistical analysis. For ADAPT-MS classification of validation samples, no imputation was applied; the framework uses only the proteins detected in each individual sample.

### Statistical analysis

All processing, statistical analysis and visualization were performed in Python 3.12.11 using numpy 2.3.5, pandas 2.3.3, scipy 1.16.3, statsmodels 0.14.6, scikit-learn 1.7.2, matplotlib 3.10.7, seaborn 0.13.2,networkx 3.6.1, gseapy, pyComBat 0.3.3 and adjustText 1.3.0. All tests were two-sided, and false discovery rate control was applied within each analysis. No formal sample size calculation was performed; all available samples that met the eligibility criteria were analyzed.

Differential abundance across the four diagnostic categories was assessed by one-way ANOVA with Benjamini–Hochberg (BH) false-discovery-rate control; pairwise category comparisons used Welch t-tests on non-imputed data with missing values omitted per test, with BH control and q<0.01 were considered significant unless stated otherwise. Significant proteins were clustered by Ward linkage on Euclidean distance, and Gene Ontology biological-process over-representation of each cluster was tested with Enrichr through gseapy (BH-adjusted). Fold changes of the benign-versus-malignant and benign-versus-metastasis-to-ovary comparisons were compared by Pearson correlation. Biological-subgroup analyses compared epithelial and non-epithelial benign masses with their malignant counterparts using the same pipeline.

To characterize metadata structure, proteins significantly associated (q<0.01) with sample category, age, race, menopausal status or body mass index (BMI, ANOVA for categorical, Spearman correlation for continuous parameters) were assembled into a Spearman co-expression network (|r|≥0.55). The network was partitioned into modules by Louvain community detection, retaining modules with at least ten proteins; module identities were annotated by over-representation analysis through the STRING enrichment interface, and module extents were drawn as convex hulls. Associations between individual proteins and continuous covariates were displayed as LOWESS fits with bootstrap confidence bands. To assess confounding of the benign-versus-malignant contrast, each significant protein was modeled by ordinary least squares as protein ∼ malignancy + age + BMI + race + menopause. Variance was partitioned among terms by Type-II ANOVA and expressed as partial η^2^, and the malignancy term was re-tested after adjustment with BH control; proteins whose largest covariate partial η^2^ exceeded that of malignancy were flagged as covariate-dominated.

Classifier discrimination was summarized by the area under the receiver operating characteristic (ROC) curve, computed for each validation cohort with 95% confidence intervals from 2000 bootstrap resamples. Because the five cohorts constitute independent replications and differ in prevalence and score distribution, cohort-specific AUCs were combined by DerSimonian–Laird random-effects meta-analysis with variances from DeLong’s method rather than by pooling samples, which inflates apparent discrimination through between-cohort differences; between-cohort heterogeneity is reported as I^2^, the percentage of variance not attributable to sampling error. Sensitivity, specificity, positive and negative predictive value, and accuracy are reported pooled at a probability threshold of 0.5 for ADAPT-MS and 35 U/mL for serum CA-125. Held-out discovery performance, obtained by the five-fold cross-validation described in the ADAPT-MS section, was compared with validation performance using an unpaired z-test of the difference between the two summary AUCs. The confidence interval of that difference is reported, since a non-significant difference alone would not establish equivalence. Reclassification between models was quantified as the number of samples crossing the 0.5 threshold in each direction.

Single-protein comparators were evaluated on the single-sample protein matrices with the direction of effect fixed from the discovery cohort and are reported with bootstrap confidence intervals along with the fraction of samples in which each protein was quantified, since coverage differs markedly between markers. Protein identification depth between the discovery and validation cohorts was compared by the Mann-Whitney U test.

Clinical utility was assessed by decision-curve analysis, reporting net benefit across threshold probabilities relative to treat-all and treat-none strategies, and by calibration of predicted against observed probabilities; Youden-optimal and 95%-sensitivity operating points are reported. Classifier robustness was examined in prespecified subgroups (menopausal status, germline BRCA1/2 or Lynch-syndrome carrier status, age 50–70 years, body-mass index above and below 25 kg/m^2^, and FIGO (2013) stage) with AUCs and bootstrap confidence intervals computed on samples pooled across validation cohorts, because individual subgroups are too small for cohort-level estimation.

Sensitivity of the classifier output to clinical and technical characteristics was assessed separately within each diagnostic class, where the true label is constant so that any association with the predicted probability of malignancy represents a prediction bias rather than a causal effect. Continuous variables were assessed using Spearman correlation with LOWESS fits, and categorical variables by Kruskal-Wallis tests with pairwise Mann-Whitney comparisons for the clinically and equity-relevant contrasts. Effects were additionally estimated with both classes pooled and class included as a covariate, and then with validation cohort added as a covariate, since identification depth and contamination vary systematically between cohorts; whether discrimination itself degraded was tested by computing AUC within tertiles of each variable. Technical variables comprised the number of quantified protein groups per sample and four contamination indices — erythrocyte, platelet, peripheral blood mononuclear cell (PBMC) and coagulation — each defined as the summed raw intensity of its marker panel divided by the summed raw intensity of all proteins quantified in that sample, computed from the single-sample protein matrices so that they describe the data the classifier actually receives. Erythrocyte, platelet, and PBMC panels were taken from the published PCA-N marker sets (Korff et al. 2025); the coagulation panel used blood-coagulation quality markers (Geyer et al. 2019), which by design include platelet-derived proteins. Because incomplete clotting releases platelet contents, the index is expected to correlate with the platelet index and their correlation is reported. When a gene mapped to more than one protein group, the best-quantified group was used to avoid double-counting a marker.

### ADAPT-MS classification framework

We applied ADAPT-MS (Müller-Reif et al. 2026), with refinements to feature handling. The classification task was binary and restricted to adnexal masses: the discovery training set comprised 485 samples (123 malignant, including tumors metastatic to ovary (MetToOv), and 362 benign adnexal masses); healthy controls (n=577) were not used for classifier training or evaluation.

A relaxed feature list was generated from the discovery cohort using Welch t-test feature selection across ten random stratified 80/20 splits on non-imputed data (missing values omitted from each test, unequal variances assumed). Within each split, p-values were adjusted by the Benjamini–Hochberg procedure and the top-N features by adjusted q-value were retained. The relaxed list is the union across the ten splits, thus combining the top-N features of all folds, to make feature selection more robust. The number of top features per split was tuned in a sensitivity analysis over 5 to 200 features; 125 maximized validation performance for the sequential classifier and 75 for the Basic classifier, giving relaxed lists of approximately 175 features.

Each validation sample was then classified independently. For that sample, the intersection of the relaxed list with the proteins actually quantified in it was formed. Importantly, undetected proteins were not zero-filled, and the validation sample was never imputed, as the ADAPT-MS procedure uses only proteins per sample that are actually quantified. An ElasticNet logistic regression model (C=1.0, L1 ratio 0.9, saga solver) was refitted on the discovery data restricted to those features, with k-nearest-neighbor imputation (k=5) applied only to that discovery sub-matrix. Samples with fewer than five available features would have been left unclassified; no sample met this criterion (0 of 623). The classifier returns a probability of malignancy for each sample, at a >=0.5 threshold where a binary call is required.

We extended the base framework with a sequential learning strategy in which each classified validation sample was appended to the training set, along with its pathology-confirmed (true) label before the next sample was classified, with the relaxed feature list refreshed every ten added samples. Cohorts were processed in order Val1, Val2, Val3, Val4, and the external cohort ValRP, and within each cohort in the order in which raw files were enumerated; because the sequential configuration accumulates labels, results depend on this order. This configuration leverages the surgical feedback loop inherent to the treatment of women with adnexal masses, in which the pathology-confirmed diagnosis of each patient undergoing surgery becomes a verified training label for subsequent cases. The non-sequential Basic classifier, trained only on the discovery cohort, serves as the static-model reference and represents the lower bound for realistic deployment.

To estimate internal performance on held-out data, the discovery cohort was additionally analyzed by stratified five-fold cross-validation. Within every fold, the classifier was constructed on the training folds alone, so that feature selection never saw held-out samples; the held-out samples were then classified one at a time through the identical per-sample procedure described above, retaining their original missingness so that each presented a variable feature space as a single-sample validation search does. Feature selection performed once on the full discovery cohort before cross-validation would have leaked and inflated this estimate. Splits used seeds 0–9 for feature selection and a fixed seed of 42 for model fitting, imputation and cross-validation, so all reported values are deterministic.

### Integration with O-RADS ultrasound and plasma biomarkers

For a subset of validation patients, pre-operative Ovarian-Adnexal Reporting and Data System (O-RADS) (version 2) (Yoeli-Bik et al. 2023; Strachowski et al. 2023) ultrasound scores and serum CA-125 levels measured within one week of blood collection were available. Sonographic evaluations were conducted at the University of Chicago Medical Center, Department of Obstetrics and Gynecology, using GE HealthCare Voluson E8 and E10 and Samsung Elite WS80 ultrasound machines. Ultrasound was performed by experienced sonographers and systematically reviewed by a board-certified OB&GYN (AR, MS), with a consensus of expert US examiners in about 20% of cases, providing an audit of the accuracy and quality of imaging assessment using all available images, including cine clips. If a patient had multiple adnexal masses, the O-RADS score was determined by the most suspicious characteristics and used in the study.

To assess complementarity, we performed matched 1:1 comparisons: for each external predictor, we restricted the analysis to patients for whom that predictor was available and compared its discrimination with the ADAPT-MS proteomic classifier evaluated on exactly those patients. For the combined model, the pre-operative O-RADS score was appended as an additional feature on its ordinal 1–5 scale within the same sequential ElasticNet framework and hyperparameters, with training at each step restricted to samples carrying a recorded O-RADS score. The comparators were: O-RADS alone; serum CA-125 alone, used as a continuous value for discrimination at a threshold of 35 U/mL for binary metrics; a proteome-derived ROMA-like model (proteomic CA-125 and HE4 with menopausal status and age); a two-marker proteomic model (CA-125 and HE4 only); and single-protein classifiers (proteomic CA-125 and HE4 alone). The ROMA-like and two-marker models used the same ElasticNet logistic regression as the full classifier, differing only in their feature sets. Single-protein classifiers were evaluated on the single-sample protein matrices, with the direction of effect fixed from the discovery cohort rather than fitted on validation data. The proteomic reference predictions for these comparisons were taken from the sequential ADAPT-MS classifier. Feature availability across cohorts is summarized in Supplementary Figure 8A.

## Data Availability

All data produced in the present study are available upon reasonable request to the authors.

## AUTHOR CONTRIBUTIONS

JBMR: conceptualization, data curation, investigation, formal analysis, methodology, supervision, writing (original draft writing, review, and editing), software, visualization. SA: conceptualization, data curation, investigation, formal analysis, methodology, writing (original draft writing, review, and editing). VA: data curation, investigation, formal analysis, writing (review and editing). MW: data curation, writing (review and editing). KK: data curation, formal analysis, writing (review and editing). ASS: data curation, formal analysis, writing (review and editing). RYB: data curation. AR: data curation, formal analysis. MS: data curation, formal analysis. EZ: data curation, funding acquisition, investigation, methodology, project administration, supervision. MM: conceptualization, funding acquisition, project administration, supervision, writing (original draft writing, review, and editing). EL: conceptualization, data curation, funding acquisition, investigation, methodology, project administration, supervision, writing (original draft writing, review, and editing).

## ACKNOWLEDGMENTS

We want to thank Elena Nevins, Kiersten Hogan, and Calla O’Connor from the University of Chicago, who enrolled patients and performed the plasma collection, processing, and clinical data collection. We thank Igor Paron and Tim Heymann at the Max Planck Institute of Biochemistry for assistance with LC-MS.

## SUPPLEMENTARY DATA

**Supplementary Figure 1.**
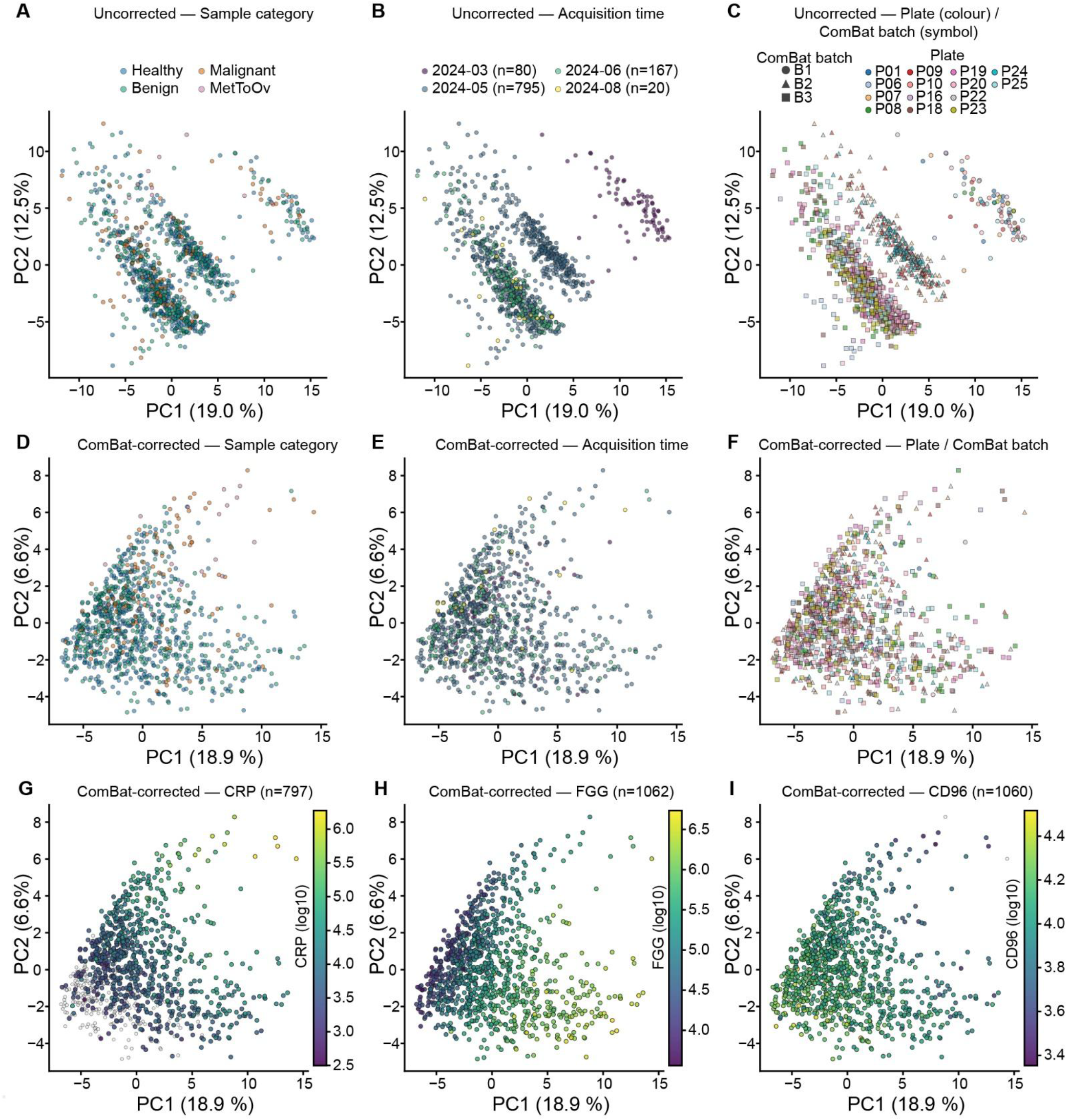
Quality control of batch-correction by principal-component analysis of the discovery proteome. Principal components 1 and 2 of all 1,062 discovery samples before (A–C) and after (D–F) pyComBat batch correction, colored three ways. (A, D) By sample category (healthy control, benign adnexal mass, malignant/borderline adnexal mass, metastasis to ovary). (B, E) By acquisition session — four mass-spectrometry runs between March and August 2024 (2024-03-03 n=80; 2024-05-29 n=795; 2024-06-12 n=167; 2024-08-21 n=20), ordered earliest to latest. (C, F) By acquisition plate (14 plates), with marker shape giving the acquisition batch (B1/B2/B3) used as the ComBat batch variable, a grouping of plate codes distinct from the plate identifier. Before correction samples separate by acquisition session and batch (B, C); afterwards this structure is removed (E, F) while the diagnostic-category structure is retained (A versus D). (G–I) Batch-corrected principal components colored by the log10 plasma intensity of CRP (G), FGG (H) and CD96 (I); grey points are samples in which the protein was not detected.

**Supplementary Figure 2.**
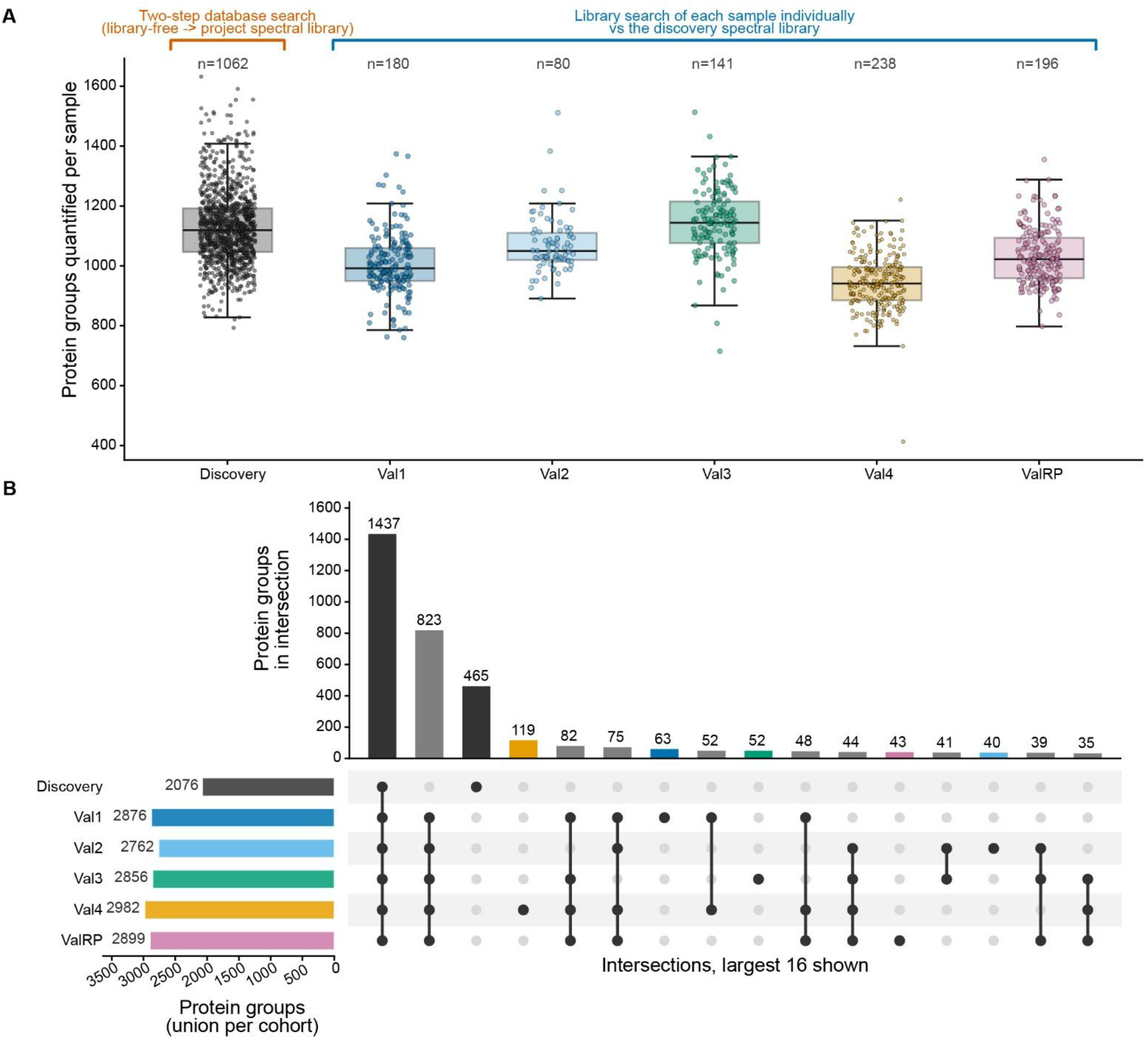
Protein groups quantified per sample and overlap across the six cohorts. (A) Protein groups quantified per sample, shown as box plots with all individual samples overlaid. Brackets above the panel mark the two search strategies: the discovery cohort was searched in two steps, library-free followed by re-search against the project spectral library, whereas each validation sample was searched individually against that discovery library — the single-sample scenario ADAPT-MS is designed for, and the reason validation samples quantify fewer proteins per run. Discovery counts are taken from the working analysis matrix, after removal of protein groups quantified in fewer than 4% of runs. (B) UpSet plot of protein-group overlap across cohorts: bars above give the size of each membership pattern (the largest 16 of 59 shown), the matrix below indicates which cohorts each pattern comprises, and horizontal bars on the left give the cumulative protein groups per cohort. Of a union of 3,941 protein groups, 1,437 are shared by all six cohorts. The discovery set is smaller than each validation union because it carries the 4% completeness filter, whereas validation unions accumulate every protein group seen in any single-sample search. Despite independent sampling and processing, the Roswell Park cohort contributes only 43 unique protein groups and identifies no shifted subset.

**Supplementary Figure 3.**
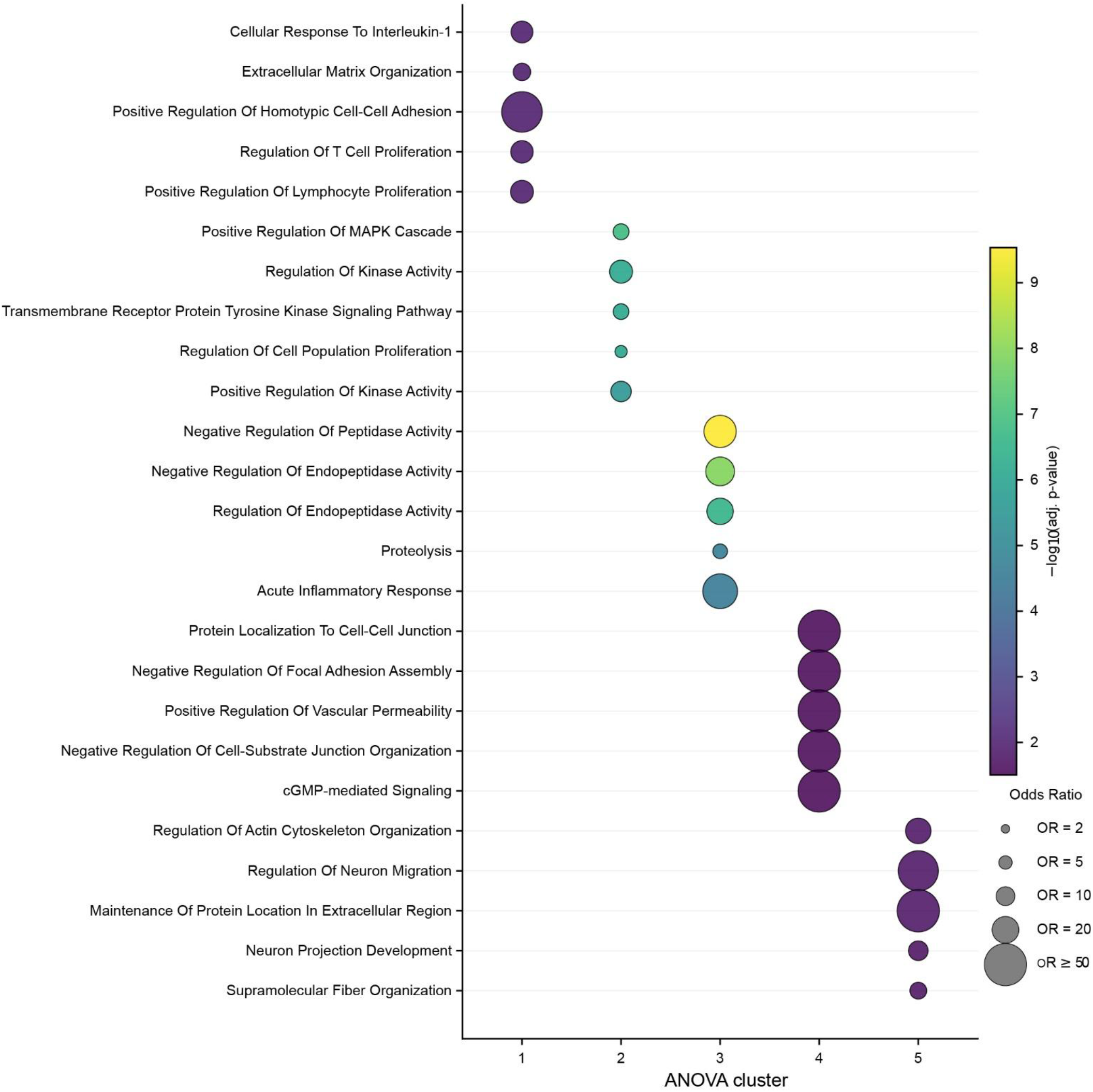
Gene-set enrichment within ANOVA-defined clusters. Over-representation of GO Biological Process terms (GO_Biological_Process_2023, tested with Enrichr) for each of the five clusters of ANOVA-significant proteins from Figure 2A. For each cluster the five most significant terms are shown, ranked by Benjamini–Hochberg-adjusted p-value; only terms at FDR<0.05 are considered. Bubble size is proportional to the odds ratio, capped at 50 for display, and color gives −log10 of the adjusted p-value.

**Supplementary Figure 4.**
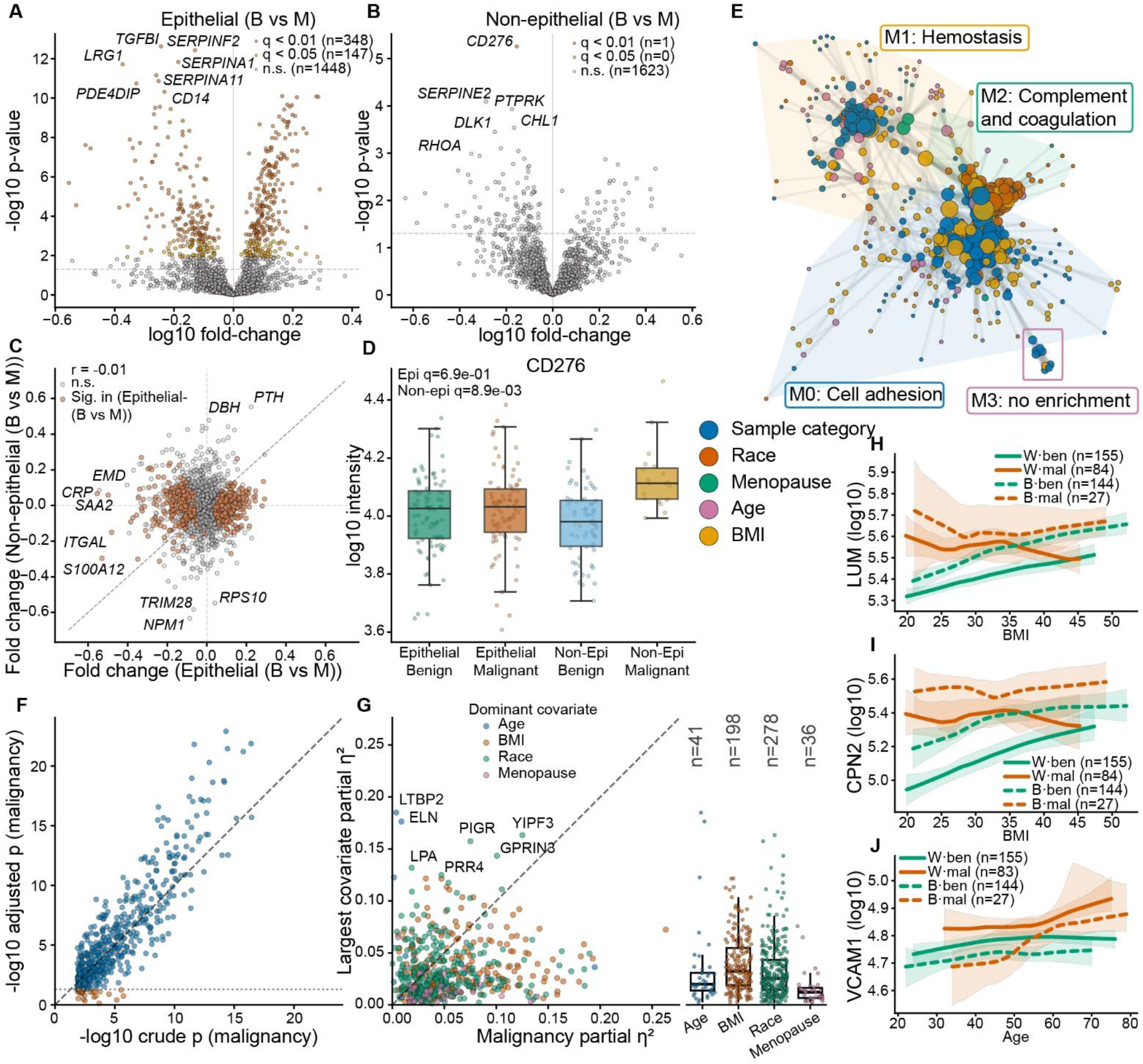
Biological subgroups and metadata structure of the discovery plasma proteome. Discovery cohort, n=1,062. (A) Volcano plot, benign versus malignant epithelial tumors (85 versus 85): 495 proteins at q<0.05. (B) The same contrast in the non-epithelial lineage (66 versus 19): one significant protein. (C) Fold changes of the epithelial (x-axis) against the non-epithelial (y-axis) comparison, colored by significance; ten largest effects labeled. (D) CD276 across the four biological subgroups of Epithelial, Non-epithelial and thereof benign and malignant individuals. (E) Co-expression network of the 505 proteins significantly associated (q<0.01) with diagnostic category, race, menopausal status, age or body-mass index. Edges are Spearman correlations (|r|≥0.55, width ∝|r|), node color the dominant metadata parameter, node size the number of connections. Louvain community detection gives four modules, shown as shaded hulls: M0 cell adhesion (GO-BP), M1 hemostasis (Reactome), M2 complement and coagulation cascades (KEGG), M3 no significant enrichment. (F) The 553 proteins, significantly discriminating benign from malignant masses (q<0.05), before (x-axis) versus after (y-axis) adjustment by ordinary least squares model for age, body-mass index, race and menopausal status, as −log10 p. The 30 proteins losing significance (adjusted p≥0.05) are vermillion; dotted line, p=0.05. (G) Malignancy partial η^2^ against the largest covariate partial η^2^ for the same proteins, from protein ∼ malignancy + age + BMI + race + menopause, colored by dominant covariate; the 167 points above the diagonal are covariate-dominated. The swarm shares the y-axis and splits it by dominant covariate (race n=278, BMI n=198, age n=41, menopause n=36). (H–J) LUM (H) and CPN2 (I) against body-mass index, VCAM1 (J) against age, with LOWESS fits and 95% confidence bands per race × diagnosis group (White solid, Black dashed; benign green, malignant vermillion).

**Supplementary Figure 5.**
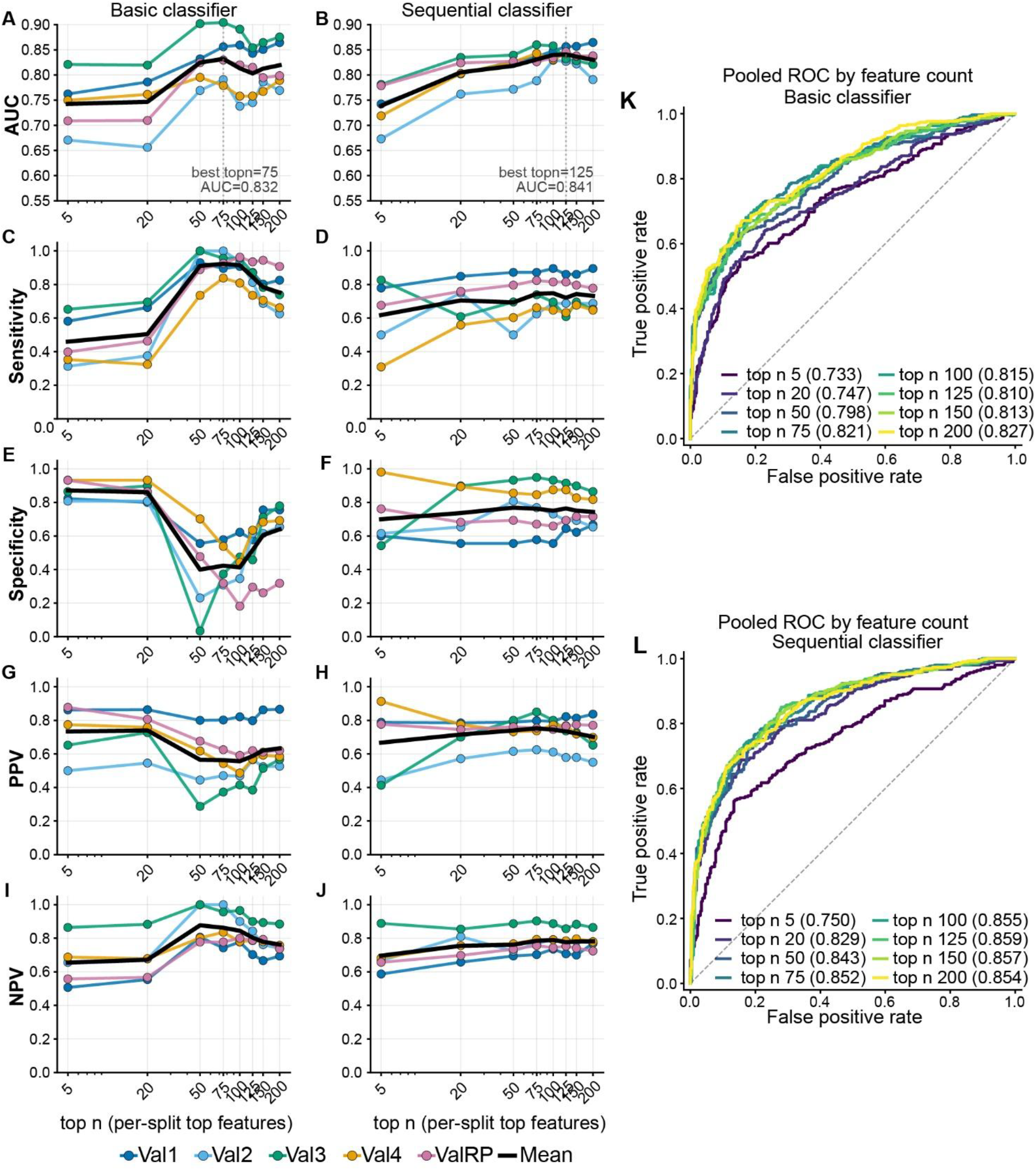
Feature-count sensitivity of ADAPT-MS. (A–J) AUC, sensitivity, specificity, PPV and NPV (columns) for the Basic (A,C,E,G,I) and sequential (B,D,F,H,J) classifiers as the number of top-ranked features selected per feature-selection split is varied over 5, 20, 50, 75, 100, 125, 150 and 200, shown for each validation cohort and as the cohort mean. Mean validation AUC peaked at 125 features for the sequential classifier (0.841) and at 75 for the Basic classifier (0.832) and was within 0.02 of its maximum from about 50 features onward in both configurations; these values were adopted for all other analyses. (K, L) Receiver-operating-characteristic curves at each feature count, pooled across validation cohorts, for the Basic (K) and sequential (L) classifiers.

**Supplementary Figure 6.**
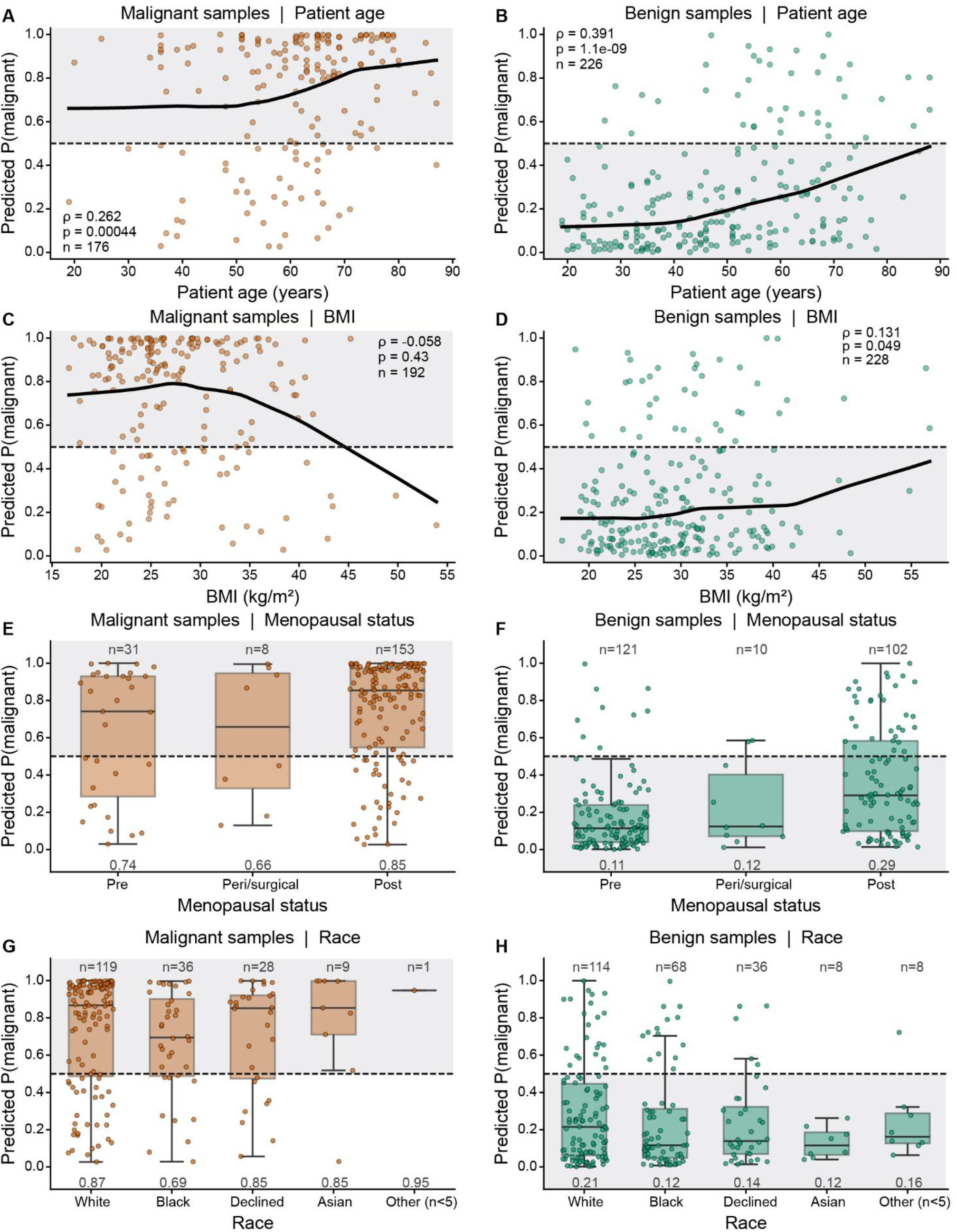
Confounder and subgroup sensitivity analyses. Predictive score from the sequential classifier is correlated separately for malignant and benign individuals against patient age (A+B), BMI (C+D), Menopausal status (E+F) and Race (G+H). For malignant samples, all points which fall above the cutoff of 0.5 are classified correctly, for benign samples, all points which fall below the cutoff of 0.5 are predicted correctly; the area where correctly classified samples are is shaded in gray.

**Supplementary Figure 7.**
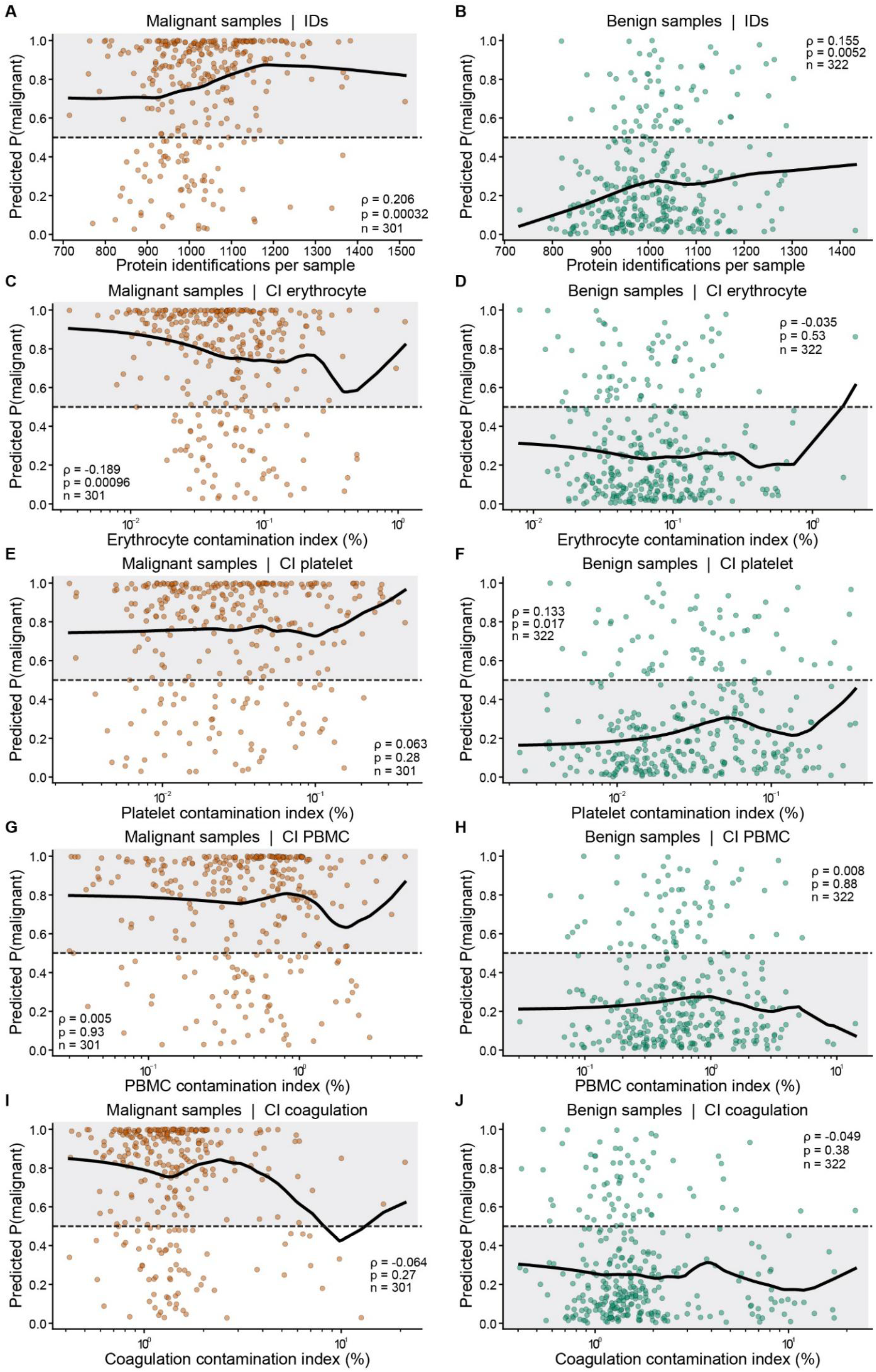
Technical and preprocessing confounder sensitivity analyses. Predictive score from the sequential classifier is correlated separately for malignant and benign individuals against number of protein identifications (A+B), Erythrocyte contamination index (C+D), Platelet contamination index (E+F), PBMC contamination index (G+H) and coagulation contamination index (I+J). For malignant samples, all points which fall above the cutoff of 0.5 are classified correctly, for benign samples, all points which fall below the cutoff of 0.5 are predicted correctly; the area where correctly classified samples are is shaded in gray.

**Supplementary Figure 8.**
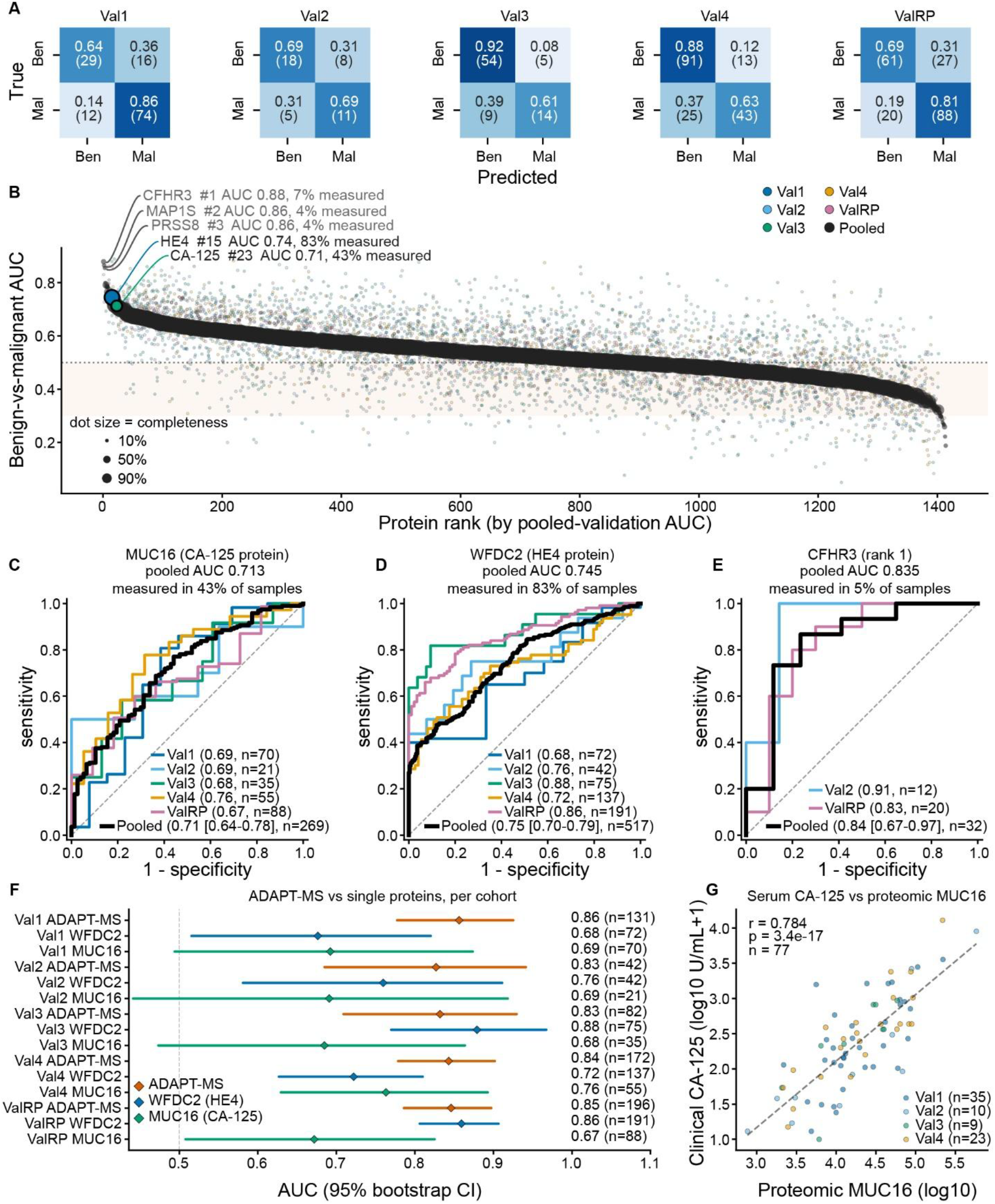
Single-protein classifiers compared with ADAPT-MS. (A) Confusion matrices per validation cohort for the sequential ADAPT-MS classifier at a malignancy-probability threshold of 0.5. (B) All 1,414 protein groups measurable in both discovery and validation, each used as a single-protein classifier and ranked by pooled validation AUC (x-axis rank, y-axis AUC), with per-cohort performance in separate colors and the three top-ranked proteins labeled with their AUC and the fraction of samples in which they were quantified: CFHR3 (0.879, 7%), MAP1S (0.865, 4%) and PRSS8 (0.855, 4%). The direction of effect for each protein was fixed from the discovery cohort. (C–E) Discrimination of proteomic MUC16/CA-125 (C; pooled AUC 0.713, measured in 43% of samples), WFDC2/HE4 (D; 0.745, 83%) and CFHR3 (E; 0.835, 5%) across validation cohorts, shown per cohort and pooled. (F) ADAPT-MS against the single-protein classifiers within each validation cohort. (G) Clinical serum CA-125 measured by ELISA against proteomic MUC16 on the samples with both available (Pearson r=0.784).

**Supplementary Figure 9.**
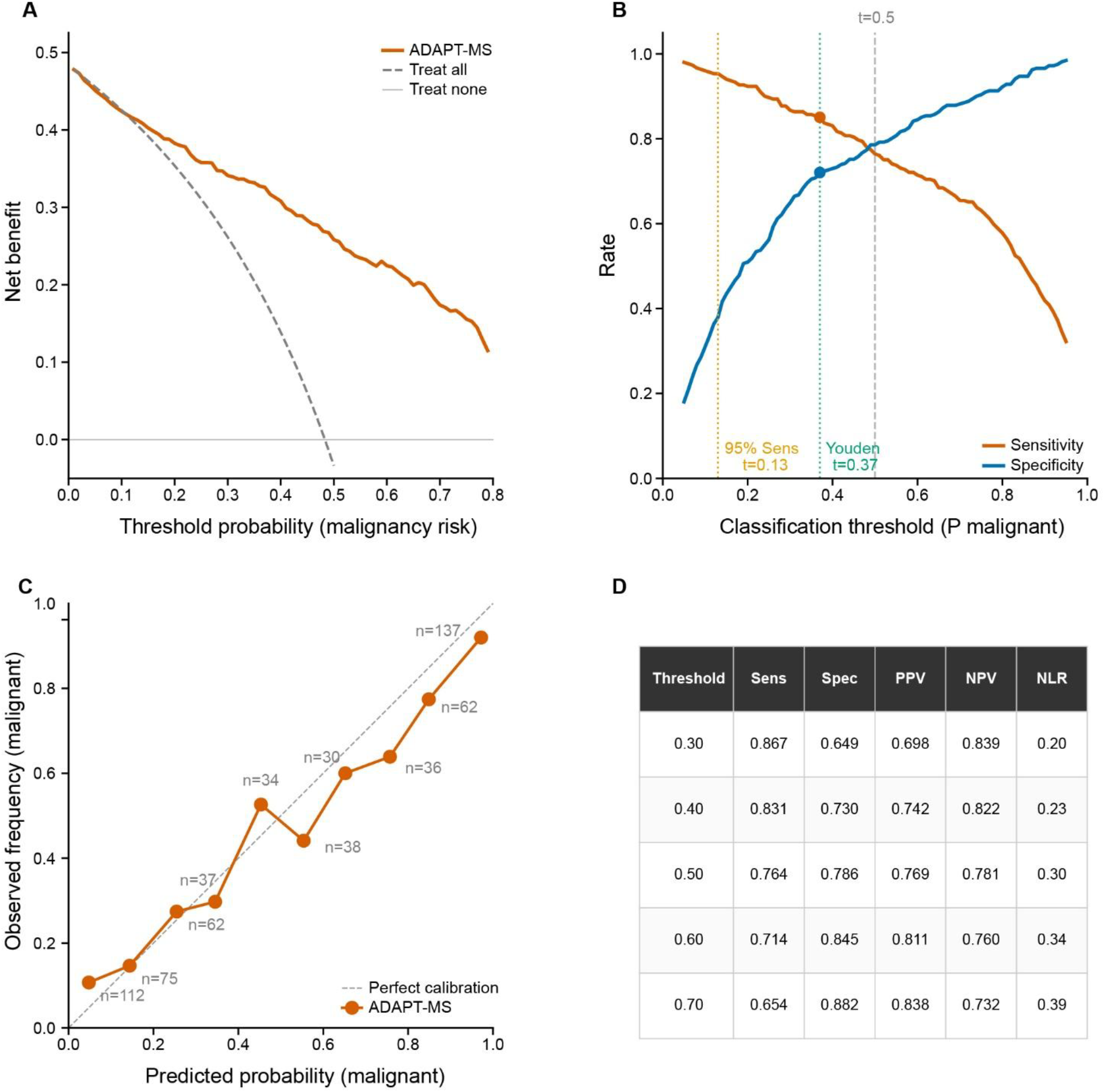
Clinical utility of ADAPT-MS with all 623 validation samples (301 malignant, 322 benign) across the five cohorts, using the sequential classifier. (A) Decision-curve analysis: Net benefit across malignancy-risk thresholds against the treat-all and treat-none strategies, with net benefit exceeding both from a threshold of 0.01 to 0.8. (B) Sensitivity and specificity across the classification threshold, marking the Youden-optimal point (t=0.37; sensitivity 0.850, specificity 0.720) and the 95%-sensitivity operating point (t=0.13; specificity 0.379). (C) Calibration of predicted against observed malignancy probability. (D) Sensitivity, specificity, PPV, NPV and negative likelihood ratio at fixed thresholds.

**Supplementary Figure 10.**
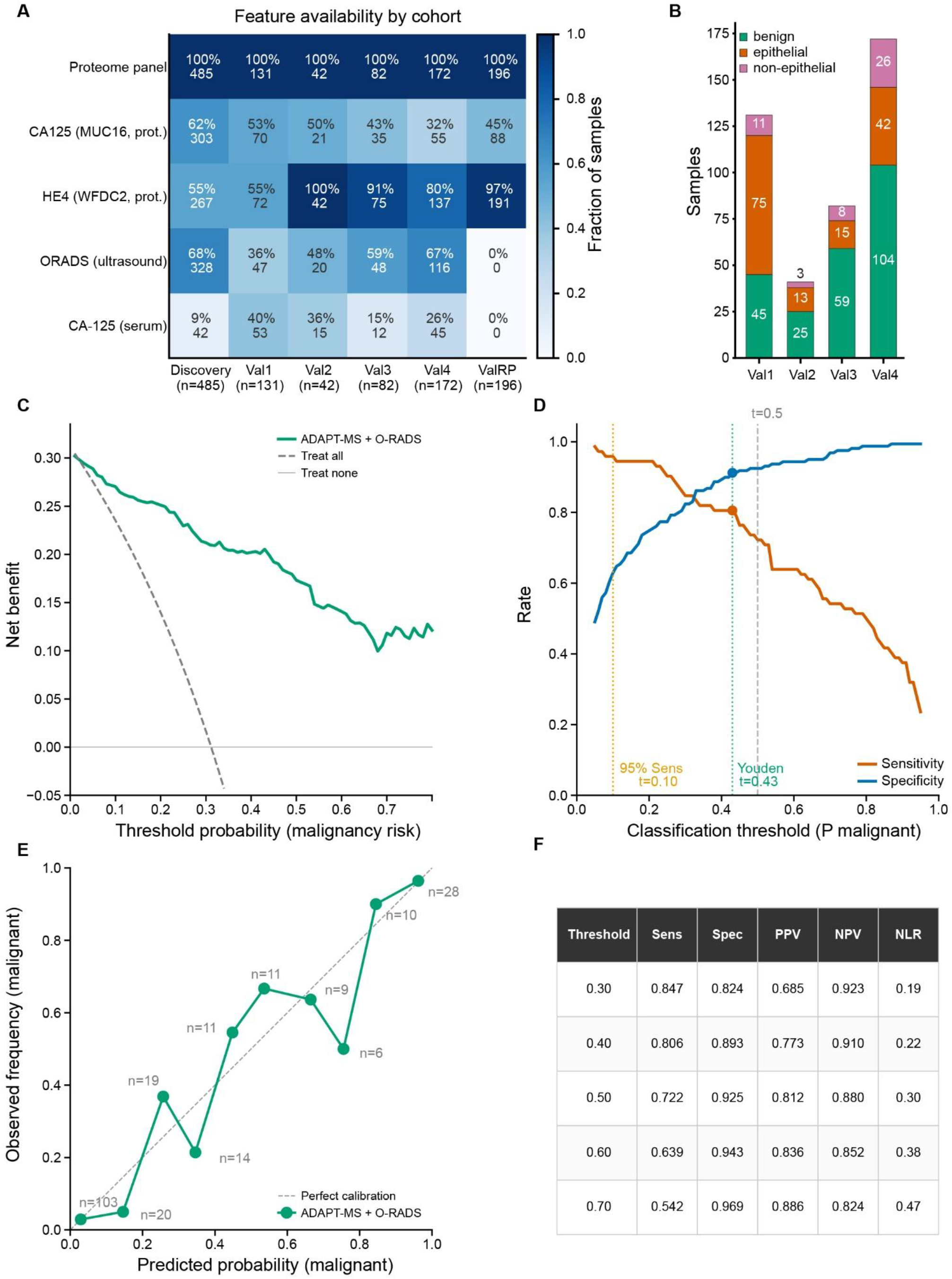
Clinical utility of the combined proteomics plus O-RADS classifier. (A) Feature-availability matrix giving, per cohort, the fraction and number of classifiable samples with each measurement: the proteome panel, proteomic MUC16 (CA-125), proteomic WFDC2 (HE4), pre-operative O-RADS ultrasound and clinical serum CA-125. (B) Composition of each validation cohort by benign, epithelial malignant and non-epithelial malignant samples. Further, the same analyses as Supplementary Figure 7, for the sequential classifier with the O-RADS ultrasound score added as a feature (Figure 4), pooled over the 231 validation samples with a pre-operative O-RADS score (72 malignant, 159 benign; Val1 47, Val2 20, Val3 48, Val4 116); the clinically applicable deployment scenario. (C) Decision-curve analysis of net benefit across malignancy-risk thresholds against treat-all and treat-none, with net benefit exceeding both from 0.02 to 0.8. (D) Sensitivity and specificity across the classification threshold, marking the Youden-optimal point (t=0.43; sensitivity 0.806, specificity 0.912) and the 95%-sensitivity operating point (t=0.10; specificity 0.629). (E) Calibration of predicted against observed malignancy probability. (F) Sensitivity, specificity, PPV, NPV and negative likelihood ratio at fixed thresholds.

**Supplementary Figure 11.**
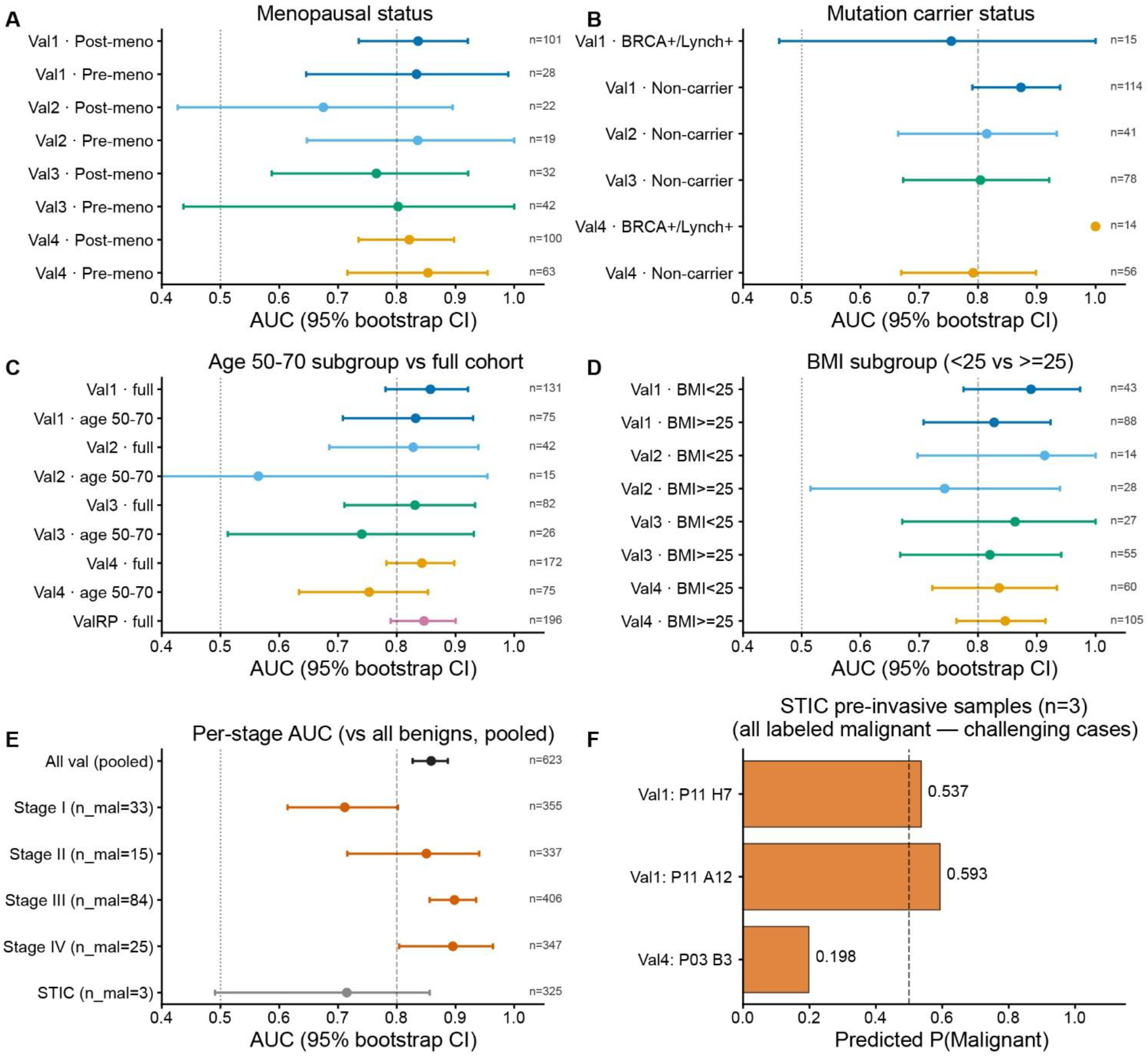
Subgroup robustness of the sequential ADAPT-MS classifier. Discrimination (AUC with 95% bootstrap confidence intervals) within prespecified clinical subgroups. (A) Menopausal status (post- versus pre-menopausal). (B) Germline mutation-carrier status (BRCA1/2 or Lynch syndrome versus non-carrier); carriers were recorded in Val1 and Val4 only. (C) Age-matched 50–70-year subgroup against the full cohort. (D) Body-mass index below versus at or above 25 kg/m^2^. (E) FIGO stage, each stage against all benign masses and pooled across cohorts; discrimination rises from stage I (0.712) to stage III (0.898) and stage IV (0.896). (F) The three STIC pre-invasive cases, shown as individual predicted probabilities of malignancy.

**Supplementary Table 2.**
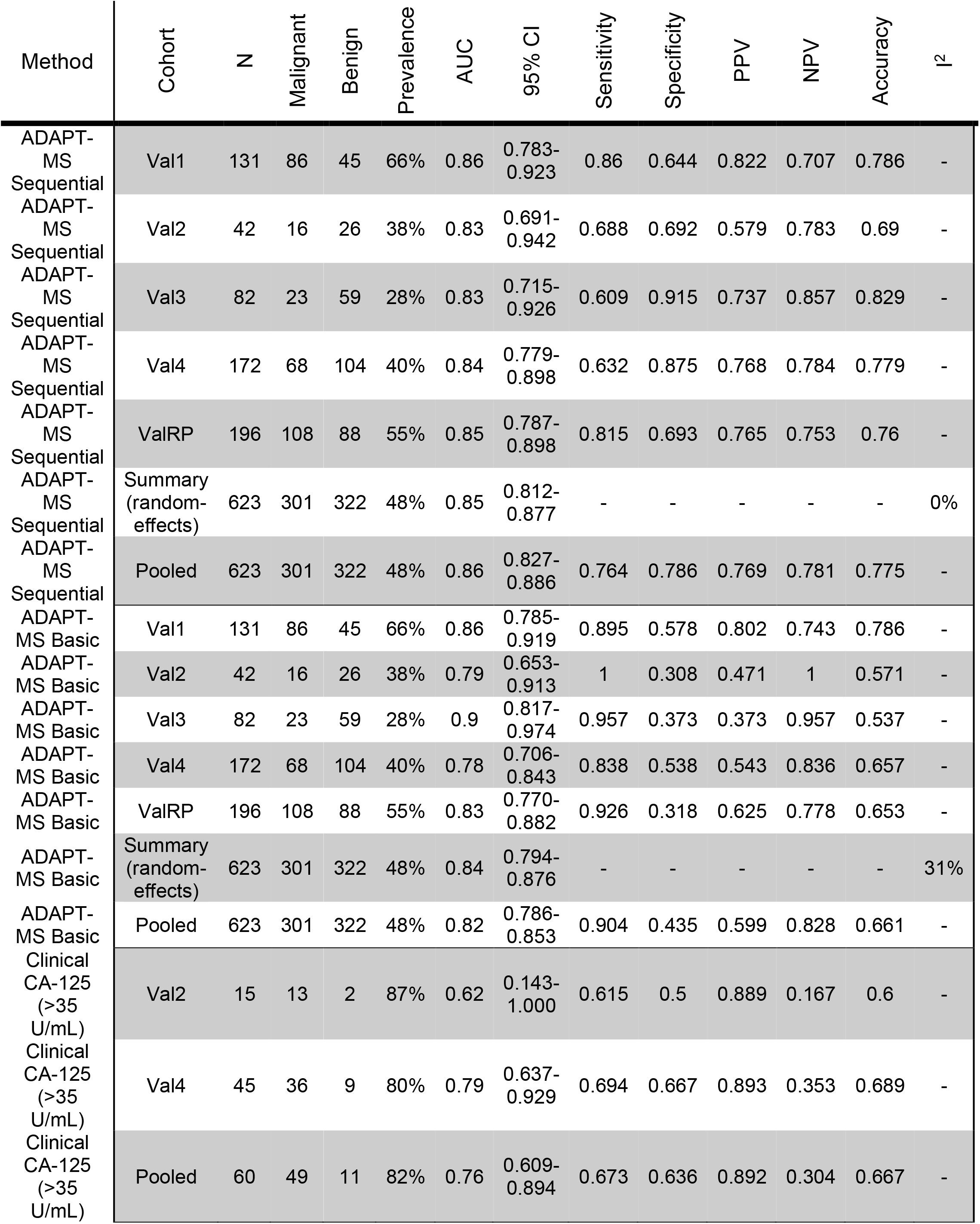
Classification performance by method and validation cohort. Scores are probability of malignancy (1 = malignant); ADAPT-MS at a threshold of 0.5 and clinical CA-125 at 35 U/mL. AUC 95% CI from 2000 bootstrap resamples for per-cohort and pooled rows. The summary row combines the cohort AUCs by DerSimonian-Laird random-effects meta-analysis (the estimate quoted in the text), with I^2^the percentage of between-cohort variance not explained by sampling error; pooling samples across cohorts inflates discrimination through between-cohort differences in score level and prevalence, so threshold-based metrics only are reported pooled. CA-125 rows cover only the samples with an available pre-operative measurement, so they are not directly comparable in number of samples to the proteomic rows.

## REFERENCES

Aebersold, R., and M. Mann. 2016. ‘Mass-spectrometric exploration of proteome structure and function’, Nature, 537: 347–55.

Albrecht, V., J. B. Müller-Reif, V. Brennsteiner, and M. Mann. 2025. ‘A Simplified Perchloric Acid Workflow With Neutralization (PCA N) for Democratizing Deep Plasma Proteomics at Population Scale’, Mol Cell Proteomics, 24: 101071.

Andreotti, R. F., D. Timmerman, L. M. Strachowski, W. Froyman, B. R. Benacerraf, G. L. Bennett, T. Bourne, D. L. Brown, B. G. Coleman, M. C. Frates, S. R. Goldstein, U. M. Hamper, M. M. Horrow, M. Hernanz-Schulman, C. Reinhold, S. L. Rose, B. P. Whitcomb, W. L. Wolfman, and P. Glanc. 2020. ‘O-RADS US Risk Stratification and Management System: A Consensus Guideline from the ACR Ovarian-Adnexal Reporting and Data System Committee’, Radiology, 294: 168–85.

Babic, A., D. W. Cramer, L. E. Kelemen, M. Köbel, H. Steed, P. M. Webb, S. E. Johnatty, A. deFazio, D. Lambrechts, M. T. Goodman, F. Heitz, K. Matsuo, S. Hosono, B. Y. Karlan, A. Jensen, S. K. Kjær, E. L. Goode, T. Pejovic, M. Moffitt, E. Høgdall, C. Høgdall, I. McNeish, and K. L. Terry. 2017. ‘Predictors of pretreatment CA125 at ovarian cancer diagnosis: a pooled analysis in the Ovarian Cancer Association Consortium’, Cancer Causes Control, 28: 459–68.

Bache, N., P. E. Geyer, D. B. Bekker-Jensen, O. Hoerning, L. Falkenby, P. V. Treit, S. Doll, I. Paron, J. B. Müller, F. Meier, J. V. Olsen, O. Vorm, and M. Mann. 2018. ‘A Novel LC System Embeds Analytes in Preformed Gradients for Rapid, Ultra-robust Proteomics’, Mol Cell Proteomics, 17: 2284–96.

Bader, J. M., V. Albrecht, and M. Mann. 2023. ‘MS-Based Proteomics of Body Fluids: The End of the Beginning’, Mol Cell Proteomics, 22: 100577.

Balkwill, F. R., C. M. Laumont, N. Burdett, O. Le Saux, D. W. Garsed, W. R. Grither, A. M. Nijhuis, M. S. Recouvreux, Z. Kang, R. Zhang, K. B. Chiappinelli, J. S. Lee, D. D. Laniti, I. McNeish, A. A. Ahmed, R. Rottapel, J. R. Conejo-Garcia, K. Odunsi, P. D. P. Pharoah, D. Zamarin, C. A. Ishak, C. Fotopoulou, R. J. Buckanovich, I. Labidi-Galy, J. D. Brenton, E. Lengyel, D. P. Cook, S. H. L. George, S. Lheureux,R. Drapkin, K. P. Nephew, S. Adams, S. Pathania, B. H. Nelson, A. Färkkilä, K. Fuh, B. G. Neel, and D. D. Bowtell. 2026. ‘Rethinking ovarian cancer III: the past decade and future directions’, Nat Rev Cancer, 26: 452–71.

Bast, R. C., Jr., T. L. Klug, E. St John, E. Jenison, J. M. Niloff, H. Lazarus, R. S. Berkowitz, T. Leavitt, C. T. Griffiths, L. Parker, V. R. Zurawski, Jr., and R. C. Knapp. 1983. ‘A radioimmunoassay using a monoclonal antibody to monitor the course of epithelial ovarian cancer’, N Engl J Med, 309: 883–7.

Bossuyt, P. M., J. B. Reitsma, D. E. Bruns, C. A. Gatsonis, P. P. Glasziou, L. Irwig, J. G. Lijmer, D. Moher, D. Rennie, H. C. de Vet, H. Y. Kressel, N. Rifai, R. M. Golub, D. G. Altman, L. Hooft, D. A. Korevaar, and J. F. Cohen. 2015. ‘STARD 2015: an updated list of essential items for reporting diagnostic accuracy studies’, Bmj, 351: h5527.

Chen, X., X. Li, X. Hu, F. Jiang, Y. Shen, R. Xu, L. Wu, P. Wei, and X. Shen. 2020. ‘LUM Expression and Its Prognostic Significance in Gastric Cancer’, Front Oncol, 10: 605.

Collins, G. S., K. G. M. Moons, P. Dhiman, R. D. Riley, A. L. Beam, B. Van Calster, M. Ghassemi, X. Liu, J. B. Reitsma, M. van Smeden, A. L. Boulesteix, J. C. Camaradou, L. A. Celi, S. Denaxas, A. K. Denniston, B. Glocker, R. M. Golub, H. Harvey, G. Heinze, M. M. Hoffman, A. P. Kengne, E. Lam, N. Lee, E. W. Loder, L. Maier-Hein, B. A. Mateen, M. D. McCradden, L. Oakden-Rayner, J. Ordish, R. Parnell, S. Rose, K. Singh, L. Wynants, and P. Logullo. 2024. ‘TRIPOD+AI statement: updated guidance for reporting clinical prediction models that use regression or machine learning methods’, Bmj, 385: e078378.

Colombo, N., C. Sessa, A. du Bois, J. Ledermann, W. G. McCluggage, I. McNeish, P. Morice, S. Pignata, I. Ray-Coquard, I. Vergote, T. Baert, I. Belaroussi, A. Dashora, S. Olbrecht, F. Planchamp, and D. Querleu. 2019. ‘ESMO-ESGO consensus conference recommendations on ovarian cancer: pathology and molecular biology, early and advanced stages, borderline tumours and recurrent disease†’, Ann Oncol, 30: 672–705.

Davenport, C., N. Rai, P. Sharma, J. J. Deeks, S. Berhane, S. Mallett, P. Saha, R. Champaneria, S. E. Bayliss, K. I. Snell, and S. Sundar. 2022. ‘Menopausal status, ultrasound and biomarker tests in combination for the diagnosis of ovarian cancer in symptomatic women’, Cochrane Database Syst Rev, 7: Cd011964.

Demichev, V., C. B. Messner, S. I. Vernardis, K. S. Lilley, and M. Ralser. 2020. ‘DIA-NN: neural networks and interference correction enable deep proteome coverage in high throughput’, Nat Methods, 17: 41–44.

Ferlay, J., M. Colombet, I. Soerjomataram, C. Mathers, D. M. Parkin, M. Piñeros, A. Znaor, and F. Bray. 2019. ‘Estimating the global cancer incidence and mortality in 2018: GLOBOCAN sources and methods’, Int J Cancer, 144: 1941–53.

Fritsche, H. A., and R. G. Bullock. 2023. ‘A reflex testing protocol using two multivariate index assays improves the risk assessment for ovarian cancer in patients with an adnexal mass’, Int J Gynaecol Obstet, 162: 485–92.

Geyer, P. E., E. Voytik, P. V. Treit, S. Doll, A. Kleinhempel, L. Niu, J. B. Müller, M. L. Buchholtz, J. M. Bader, D. Teupser, L. M. Holdt, and M. Mann. 2019. ‘Plasma Proteome Profiling to detect and avoid sample-related biases in biomarker studies’, EMBO Mol Med, 11: e10427.

Guo, T., J. A. Steen, and M. Mann. 2025. ‘Massspectrometry-based proteomics: from single cells to clinical applications’, Nature, 638: 901–11.

Guzman, U. H., A. Martinez-Val, Z. Ye, E. Damoc, T. N. Arrey, A. Pashkova, S. Renuse, E. Denisov, J. Petzoldt, A. C. Peterson, F. Harking, O. Østergaard, R. Rydbirk, S. Aznar, H. Stewart, Y. Xuan, D. Hermanson, S. Horning, C. Hock, A. Makarov, V. Zabrouskov, and J. V. Olsen. 2024. ‘Ultra-fast labelfree quantification and comprehensive proteome coverage with narrow-window data-independent acquisition’, Nat Biotechnol, 42: 1855–66.

Hendricks, N. G., S. D. Bhosale, A. J. Keoseyan, J. Ortiz, A. Stotland, S. Seyedmohammad, C. D. L. Nguyen, J. T. Bui, A. Moradian, S. M. Mockus, and J. E. Van Eyk. 2024. ‘An Inflection Point in High-Throughput Proteomics with Orbitrap Astral: Analysis of Biofluids, Cells, and Tissues’, J Proteome Res, 23: 4163–69.

Jiménez-Sánchez, A., P. Cybulska, K. L. Mager, S. Koplev, O. Cast, D. L. Couturier, D. Memon, P. Selenica, I. Nikolovski, Y. Mazaheri, Y. Bykov, F. C. Geyer, G. Macintyre, L. M. Gavarró, R. M. Drews, M. B. Gill, A. D. Papanastasiou, R. E. Sosa, R. A. Soslow, T. Walther, R. Shen, D. S. Chi, K. J. Park, T. Hollmann, J. S. Reis-Filho, F. Markowetz, P. Beltrao, H. A. Vargas, D. Zamarin, J. D. Brenton, A. Snyder, B. Weigelt, E. Sala, and M. L. Miller. 2020. ‘Unraveling tumor-immune heterogeneity in advanced ovarian cancer uncovers immunogenic effect of chemotherapy’, Nat Genet, 52: 582–93.

Johnson, W. E., C. Li, and A. Rabinovic. 2007. ‘Adjusting batch effects in microarray expression data using empirical Bayes methods’, Biostatistics, 8: 118–27.

Korff, K., J. B. Müller-Reif, D. Fichtl, V. Albrecht, A. S. Schebesta, E. C. M. Itang, S. V. Winter, L. M. Holdt, D. Teupser, M. Mann, and P. E. Geyer. 2025. ‘Preanalytical drivers of bias in bead-enriched plasma proteomics’, EMBO Mol Med, 17: 3174–96.

Kurnit, K. C., G. F. Fleming, and E. Lengyel. 2021. ‘Updates and New Options in Advanced Epithelial Ovarian Cancer Treatment’, Obstet Gynecol, 137: 108–21.

Molina, R., J. M. Escudero, J. M. Augé, X. Filella, L. Foj, A. Torné, J. Lejarcegui, and J. Pahisa. 2011. ‘HE4 a novel tumour marker for ovarian cancer: comparison with CA 125 and ROMA algorithm in patients with gynaecological diseases’, Tumour Biol, 32: 1087–95.

Müller-Reif, J. B., V. Albrecht, V. Brennsteiner, J. M. Bader, P. V. Treit, N. J. Wewer Albrechtsen, S. Pangratz-Führer, and M. Mann. 2026. ‘An adaptive, continuous-learning framework for clinical decisionmaking from proteome-wide biofluid data’, Nat Commun, 17: 1105.

NCI. 2020. “Cell-Free DNA: Biospecimen Collection and Processing. NCI Biospecimen Evidence-Based Practices.” In.

NCI. 2026 (4th Edition). “NCI Best Practices for Biospecimen Resources.” In.: Biorepositories and Biospecimen Research Branch, National Institutes of Health, U.S. Department of Health and Human Services.

‘Practice Bulletin No. 174: Evaluation and Management of Adnexal Masses’. 2016. Obstet Gynecol, 128: e210–e26.

Ricciuti, J., Q. Liu, Anmnh Khan, J. M. Joseph, B. Veuskens, T. Giridharan, S. Suzuki, T. Emmons, M. Yaffe, T. W. Kuijpers, I. Jongerius, M. Brouwer, R. B. Pouw, K. Odunsi, P. Frederick, K. L. Mager, S. Lele, N. Gaulin, C. Hakim, R. P. Edwards, A. B. Olawaiye, P. Sukamanovich, S. Taylor, E. Elishaev, E. Zsiros, F. Modugno, K. Moysich, and B. Segal. 2025. ‘Prognostic significance of serum complement activation, neutrophil extracellular traps and extracellular DNA in newly diagnosed epithelial ovarian cancer’, Gynecol Oncol, 193: 49–57.

Rifai, N., M. A. Gillette, and S. A. Carr. 2006. ‘Protein biomarker discovery and validation: the long and uncertain path to clinical utility’, Nat Biotechnol, 24: 971–83.

Siegel, R. L., T. B. Kratzer, N. S. Wagle, H. Sung, and A. Jemal. 2026. ‘Cancer statistics, 2026’, CA Cancer J Clin, 76: e70043.

Stewart, H., D. Grinfeld, J. Petzoldt, B. Hagedorn, M. Skoblin, A. Makarov, and C. Hock. 2024. ‘Crowd control of ions in the Astral analyzer’, J Mass Spectrom, 59: e5006.

Strachowski, L. M., P. Jha, C. H. Phillips, M. M. Blanchette Porter, W. Froyman, P. Glanc, Y. Guo, M. D. Patel, C. Reinhold, E. J. Suh-Burgmann, D. Timmerman, and R. F. Andreotti. 2023. ‘O-RADS US v2022: An Update from the American College of Radiology’s Ovarian-Adnexal Reporting and Data System US Committee’, Radiology, 308: e230685.

Ueland, F. R., C. P. Desimone, L. G. Seamon, R. A. Miller, S. Goodrich, I. Podzielinski, L. Sokoll, A. Smith, J. R. van Nagell, Jr., and Z. Zhang. 2011. ‘Effectiveness of a multivariate index assay in the preoperative assessment of ovarian tumors’, Obstet Gynecol, 117: 1289–97.

Viode, A., P. van Zalm, K. K. Smolen, B. Fatou, D. Stevenson, M. Jha, O. Levy, J. Steen, and H. Steen. 2023. ‘A simple, time- and cost-effective, high-throughput depletion strategy for deep plasma proteomics’, Sci Adv, 9: eadf9717.

Wolff, G., A. E. Taranko, I. Meln, J. Weinmann, T. Sijmonsma, S. Lerch, D. Heide, A. T. Billeter, D. Tews, D. Krunic, P. Fischer-Posovszky, B. P. Müller-Stich, S. Herzig, D. Grimm, M. Heikenwälder, W. W. Kao, and A. Vegiopoulos. 2019. ‘Diet-dependent function of the extracellular matrix proteoglycan Lumican in obesity and glucose homeostasis’, Mol Metab, 19: 97–106.

Wu, J., H. Yin, J. Zhu, R. J. Buckanovich, J. D. Thorpe, J. Dai, N. Urban, and D. M. Lubman. 2015. ‘Validation of LRG1 as a potential biomarker for detection of epithelial ovarian cancer by a blinded study’, PLoS One, 10: e0121112.

Yoeli-Bik, R., R. E. Longman, K. Wroblewski, M. Weigert, J. S. Abramowicz, and E. Lengyel. 2023. ‘Diagnostic Performance of Ultrasonography-Based Risk Models in Differentiating Between Benign and Malignant Ovarian Tumors in a US Cohort’, JAMA Netw Open, 6: e2323289.

